# Multi-ancestry meta-analysis of tobacco use disorder prioritizes novel candidate risk genes and reveals associations with numerous health outcomes

**DOI:** 10.1101/2023.03.27.23287713

**Authors:** Sylvanus Toikumo, Mariela V Jennings, Benjamin K Pham, Hyunjoon Lee, Travis T Mallard, Sevim B Bianchi, John J Meredith, Laura Vilar-Ribó, Heng Xu, Alexander S Hatoum, Emma C Johnson, Vanessa Pazdernik, Zeal Jinwala, Shreya R Pakala, Brittany S Leger, Maria Niarchou, Michael Ehinmowo, Penn Medicine BioBank, Million Veteran Program, Psychemerge Substance Use Disorder Workgroup, Greg D Jenkins, Anthony Batzler, Richard Pendegraft, Abraham A Palmer, Hang Zhou, Joanna M Biernacka, Brandon J Coombes, Joel Gelernter, Ke Xu, Dana B Hancock, Cox J Nancy, Jordan W Smoller, Lea K Davis, Amy C Justice, Henry R Kranzler, Rachel L Kember, Sandra Sanchez-Roige

## Abstract

Tobacco use disorder (**TUD**) is the most prevalent substance use disorder in the world. Genetic factors influence smoking behaviors, and although strides have been made using genome-wide association studies (**GWAS**) to identify risk variants, the majority of variants identified have been for nicotine consumption, rather than TUD. We leveraged five biobanks to perform a multi-ancestral meta-analysis of TUD (derived via electronic health records, **EHR**) in 898,680 individuals (739,895 European, 114,420 African American, 44,365 Latin American). We identified 88 independent risk loci; integration with functional genomic tools uncovered 461 potential risk genes, primarily expressed in the brain. TUD was genetically correlated with smoking and psychiatric traits from traditionally ascertained cohorts, externalizing behaviors in children, and hundreds of medical outcomes, including HIV infection, heart disease, and pain. This work furthers our biological understanding of TUD and establishes EHR as a source of phenotypic information for studying the genetics of TUD.

---

Tobacco use disorder (**TUD**) is the most prevalent substance use disorder in the world, with 85% of smokers meeting criteria for TUD (also known as nicotine dependence).^1,2^ TUD is a problematic pattern of tobacco use that leads to clinically significant impairment or distress.^2^ Nicotine dependent individuals often experience withdrawal symptoms when they stop smoking. As a result, they often have substantial difficulty quitting and continue to smoke despite negative mental, social, and medical consequences. Tobacco smoking is the leading cause of preventable death worldwide, causing 6 million annual premature deaths,^3^ and is also highly associated with other worldwide leading contributors of morbidity and mortality, including lung cancer, chronic obstructive pulmonary disease, cardiovascular disease, mood disorders, and other substance use disorders.^4–6^ Unfortunately, available preventative and treatment options for TUD have low success rates.^7^

Genetic factors influence smoking behaviors, with twin-heritability estimates ranging from ∼30-70%.^8–12^ Recently, genome-wide association studies (**GWAS**) have expanded in size (N∼2.5M) and yielded hundreds of novel loci for smoking-related behaviors (summarized in **Supplementary Table 1**), primarily for nicotine *consumption.*^13^ These GWAS have revealed pervasive pleiotropy, with Mendelian randomization (**MR**) analyses highlighting potential causal effects of regular tobacco smoking on health outcomes (e.g., cardiovascular health,^14^ cancer risk,^14^ bone mineral density^15^), numerous other substance use disorders (e.g., alcohol,^14^ cannabis^16^ and opioid use disorders^17^), and psychiatric and related conditions (e.g., major depressive disorder,^18^ suicide-related behaviors,^19^ loneliness^20^).

While these studies have been immensely successful, they have not focused on TUD itself, which consists of multiple components that begin with smoking initiation and regular use, and develop into problematic use, dependence, cessation, and relapse. As a result, relatively little is known about the specific genes that confer risk for the development of TUD and associated conditions. One of the major roadblocks to progress in identifying risk-conferring genes has been the lack of sufficiently large samples with *misuse* phenotypes. This is an important limitation because prior studies have shown that the genetic architecture of substance use is largely different from that of misuse^21–26^. The largest GWAS of nicotine dependence, comprising 58,000 European- and African-ancestry smokers, using the self-reported Fagerström Test for Nicotine Dependence (**FTND**), identified only five loci.^27^ In addition, while there have been nicotine dependence GWAS in individuals of ancestries other than European^28^ (**Supplementary Table 1** for full list), sample sizes for diverse populations have been limited (N<12K).

The use of electronic health records (**EHR**) is a relatively untapped, cost-effective strategy for characterizing smoking-related phenotypes, including TUD. EHR-defined TUD generally relies on International Classification of Disease (**ICD**) diagnostic codes, which can be aggregated into “phecodes” that require the presence of an ICD code on two or more separate visits. TUD diagnostic codes are effective identifiers of smoking status.^29^ A key consideration, and the one we examine in this study, is the utility of TUD phecodes for use in large-scale GWAS to boost power and improve our ability to identify novel loci for TUD.^29–31^ To address this question, we performed a multi-ancestral meta-analysis of TUD comprising 898,680 individuals of European (**EUR**), African American (**AA**) and Latin American (**LA**) ancestry recruited from multiple biobanks within the PsycheMERGE network^32^ (Vanderbilt University Medical Center’s biobank, **BioVU**, N_EUR_=46,905; Mass General Brigham Biobank, **MGBB**, N_EUR_=22,268; Penn Medicine BioBank, **PMBB**,^33^ N_EUR_=28,999, N_AA_=10,088; Million Veteran Program, **MVP**, N_EUR_=396,833, N_AA_=104,332, N_LA_=44,365), and combined with existing data from the UK Biobank (**UKBB**, N_EUR_=244,890), which used a less stringent definition. In secondary analyses, we further characterized the genetic architecture of TUD, examined pleiotropy with other psychiatric and medical outcomes, and harnessed the data to reveal new potential medications for treating this serious psychiatric condition.

## Results

### Cohort and Phenotype Descriptions

We included individuals from eight cohorts across five different sites (**Figure 1a** for an overview of the cohorts; **Supplementary Table 2** for sample sizes). The methods to ascertain cases were identical for seven of these cohorts. Individuals were identified as cases if they met criteria for a TUD phecode (a TUD ICD9 or ICD10 code on two or more separate visits, described in **Supplementary Table 3**); controls were screened for the absence of a TUD diagnostic code. We benchmarked the TUD-EHR definition against self-reported smoking questionnaire data and other comorbid ICD codes (**Supplementary Table 4**). Across contributing biobanks, cases were enriched for ever smokers (92-99%), with only a minor proportion (<2%) of cases self-identifying as never-smokers (**Supplementary Table 5**). In contrast, a smaller proportion of controls were ever smokers (17-56%), with a larger proportion self-identifying as never-smokers (39-73%). Attempts at smoking cessation were reported by 15-25% of controls and 65-95% of cases. Controls were comparable to cases on age and sex but reported much lower prevalences of other substance and psychiatric disorders than cases. Thus, almost all TUD cases have evidence of being either former or current smokers based on available self-report data.

**Figure 1.**
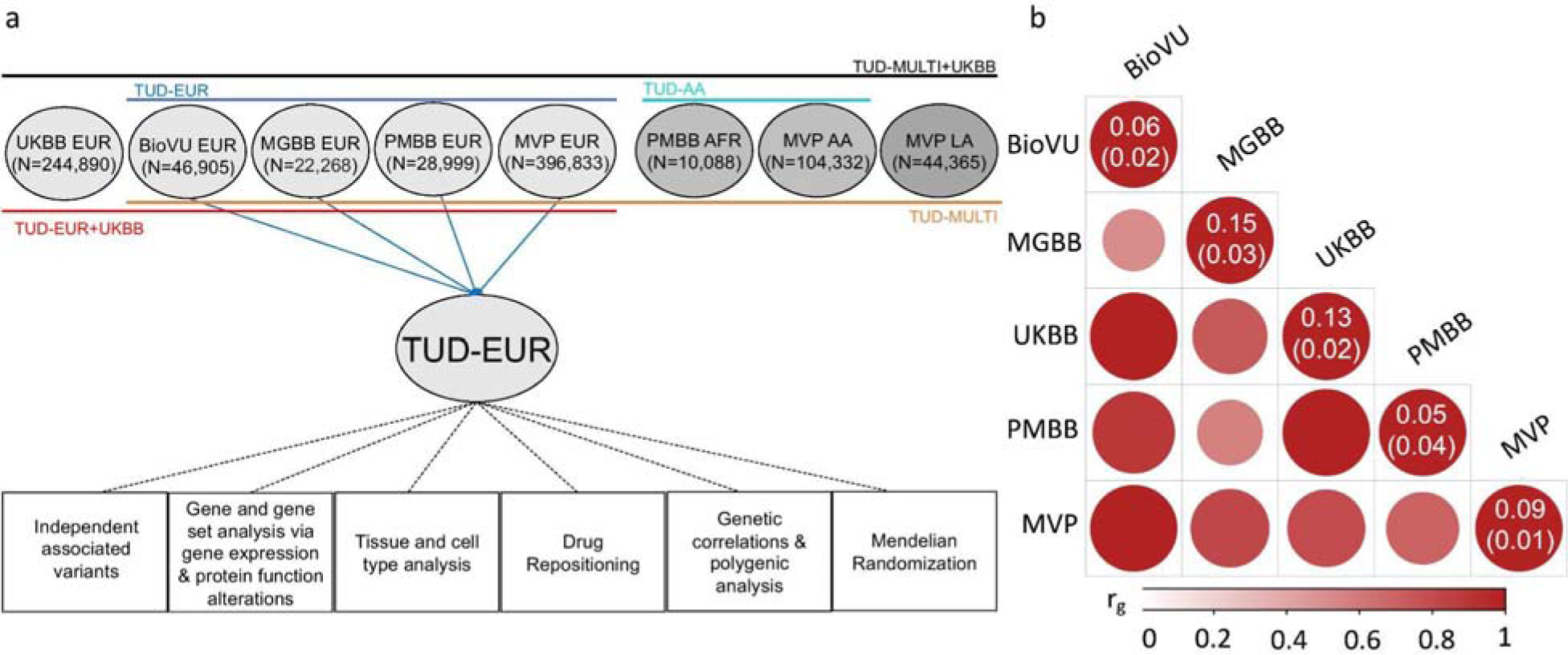
Overview of the cohorts and analysis pipeline (a) and genetic correlations among the sites (b). (**a**) We conducted independent GWAS of TUD cases and controls in individuals of European (EUR) ancestry across four PsycheMERGE sites (BioVU, MGBB, PMBB, and MVP) and performed a GWAS meta-analysis (“TUD-EUR”); these summary results were used for all secondary analyses. For African American (AA), we conducted GWAS meta-analysis of TUD cases and controls from the PMBB and MVP cohorts (“TUD-AA”). For Latin American (LA), we conducted GWAS of TUD cases and controls from the MVP cohort. Next, we performed a multi-ancestral GWAS meta-analysis (“TUD-multi”), which combined the results from all seven cohorts. We also obtained summary statistics from UKBB, which used a less stringent case definition in individuals of EUR ancestry and performed a GWAS meta-analysis within EUR individuals (“TUD-EUR+UKBB”) and across ancestries (“TUD-multi+UKBB”). **Supplementary Table 2** summarizes the datasets used for the analyses. We subjected the TUD-EUR summary statistics to several secondary analyses to characterize the genetic architecture of TUD. (**b**) LDSC genetic correlations (*r_g_*) for TUD between EUR sites were positive and high, ranging from 0.51 to unity, with most confidence intervals overlapping (**Supplementary Figure 1**). LDSC genetic correlation for TUD between the two AA samples was strongly positive (*r_g_=*0.93) but not significant (*p*=0.45). LDSC SNP-heritability estimates (*h*^2^_SNP_ 5-15%) are shown in the diagonal. UKBB=UK Biobank, BioVU=Vanderbilt University Medical Center’s biobank, MGBB=Mass General Brigham Biobank, PMBB=Penn Medicine Biobank, MVP=Million Veteran Program.

### Significant SNP-heritability and genetic correlations across sites

After applying similar data quality controls, we conducted within-cohort association analyses using logistic regression and relevant covariates (**Methods**). We estimated the proportion of variance attributable to the measured common variants (SNP-heritability, *h*^2^_SNP_) to be ∼5-15% (based on liability scale, assuming a lifetime risk of 12.5%; **Figure 1b**, **Supplementary Table 6**), which is consistent with prior nicotine-related GWAS.^13,27^ Genetic correlations across sites and ancestries were mostly high and positive (*r_g_*>0.51, *p*<1.56E-02, EUR sites; *r_g_*=0.93, *p*=0.45, AA sites; cross-ancestry *r_gs_*=0.74-0.84, *p*<3.90E-04; **Figure 1b**, **Supplementary Table 6**), serving as the basis for ancestry-specific and multi-ancestry meta-analyses, and suggesting that the genetic architecture of TUD is similar across ancestries.

### Multi-ancestry meta-analyses implicate biological underpinnings of TUD

The primary multi-ancestry meta-analysis of 20,801,211 imputed SNPs (lambda λ_GC_=1.141, **Figure 2**) was performed on seven cohorts, comprising 653,790 individuals, with 75.71% EUR, 17.50% AA, and 6.79% LA.

**Figure 2.**
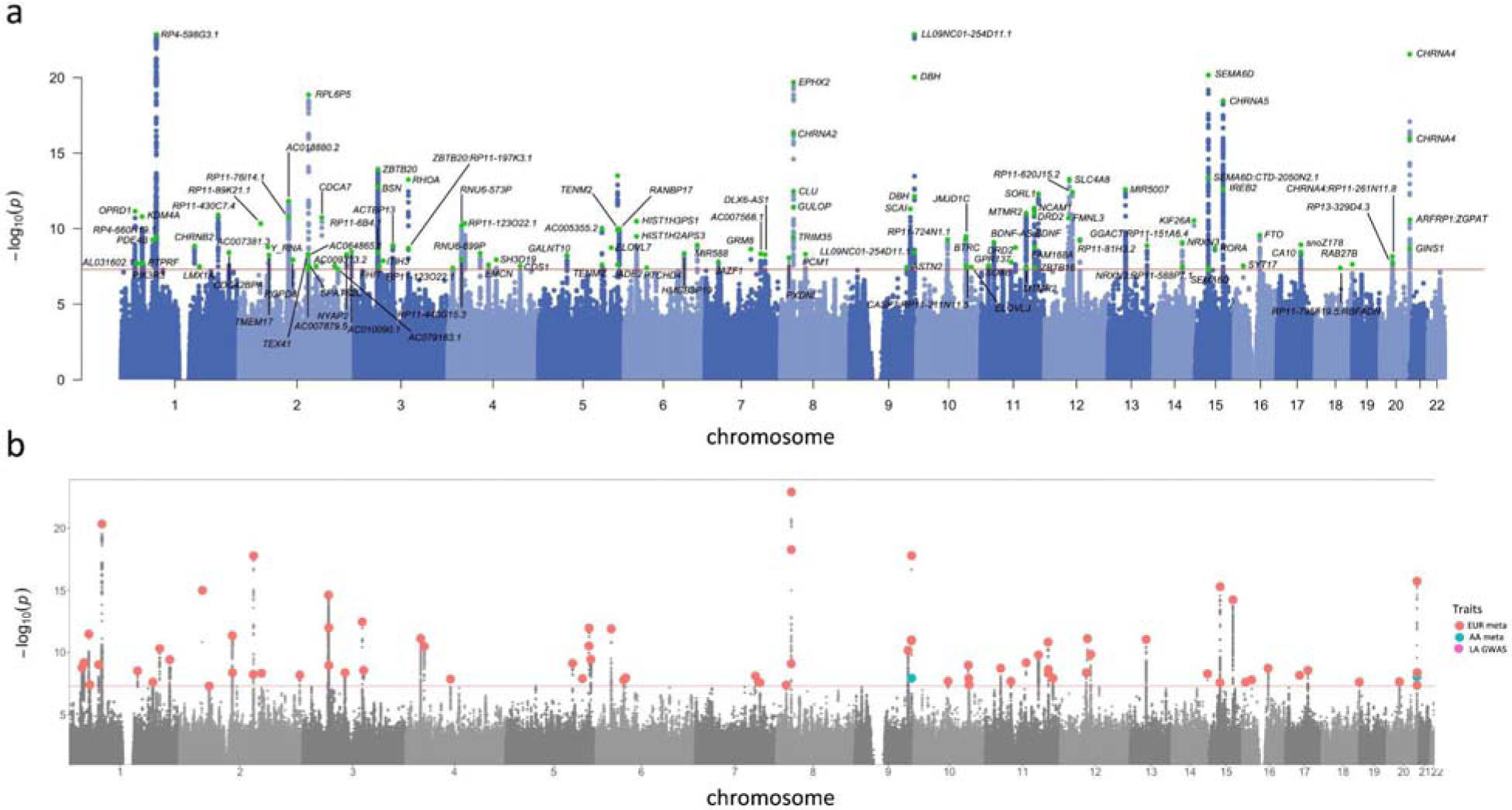
Manhattan and porcupine plots for the TUD-multi meta-analysis and ancestry-specific GWAS. (**a**) TUD-multi identified 88 independent risk loci, all of which were recently identified by the GSCAN study. (**b**) Porcupine plot of ancestry-specific meta-analyses identified 63 loci in the European cohort (EUR, in red), and 2 loci in the African-ancestry cohort (AA, in blue). No significant associations were detected in the Latin American-ancestry (LA) cohort. We used a sign test to examine the 74 EUR lead SNPs in the AA and HA cohorts, of which 57 and 53, respectively, were directly analyzed or had proxy SNPs in these populations (**Supplementary Table 10**). Most SNPs had the same direction of effect in both populations (AA = 45 out of 57, HA = 41 out of 53; sign test AA *p* =□1.31E-05, LA *p* =□8.17E-05; **Supplementary Figure 5**). Only 25 SNPs (AAs = 12, HAs = 13) were nominally associated across populations (*p* <□0.05), none of which survived multiple testing correction.

We identified 120 GWS (*p*<5.00E-08) lead SNPs (*r*^2^<0.1) located in 88 independent loci (**Supplementary Table 7**). All genome-wide significant loci had been reported by prior smoking GWAS (**Supplementary Table 7**), including aspects of smoking initiation (88/88), age of initiation (14/88), consumption (38/88), cessation (48/88) and nicotine dependence (1/88; **Supplementary Figure 2, Supplementary Figure 3**). While all these loci were recently discovered in a GWAS of 3.4 million individuals in the GSCAN study,^13^ here we reproduce some of the GSCAN findings with a considerably smaller sample size (**Supplementary Figure 3**).

Our analyses provide corroborative support for nicotinic acetylcholine receptor genes as risk genes for smoking-related traits: *CHRNA5* (rs576982, *p*=3.40E-19, chr. 15; this region includes rs16969968, a well-established functional missense polymorphism [D398N] in *CHRNA5*, *p*=2.47E-12), *CHRNB2 (*rs45490696, *p*=1.45E-09, chr. 1), *CHRNA2* (rs2741339, *p*=5.21E-17, chr. 8), and *CHRNA4* (rs2273500, *p*=2.84E-22, chr. 20). Second, we identified associations with variants in several genes that modulate dopaminergic transmission, such as the dopamine receptor D2 (*DRD2*: rs34632468, *p*=1.04E-11, and rs4936277, *p*=1.81E-09, chr.11), known for its relationship with dopamine and reward,^34^ previously associated with nicotine dependence^35^ and implicated in a recent large-scale GWAS of addiction;^36^ dopamine beta-hydroxylase (*DBH*: rs2007153, *p*=9.35E-21, and rs2519155, *p*=7.25E-13, chr.9), which encodes an enzyme necessary to convert dopamine to norepinephrine and has been consistently implicated in smoking behaviors;^13,37^ lysine demethylase 4A (*KDM4A:* rs489319, *p*=1.61E-11, chr. 1), previously found to interact with dopaminergic agents and implicated in problematic opioid use;^38^ phosphodiesterase 4B (*PDE4B*: rs7528604, *p*=5.68E-10, chr. 1), which has regulatory effects on dopaminergic pathways and has been implicated in GWAS of externalizing behaviors,^39^ smoking initiation,^37,39^ and general liability for addiction;^36^ and neural cell adhesion molecule 1, *NCAM1* (rs9919558, *p*=4.44E-12, chr. 11), which modulates dopamine signaling^41 40^ and has been associated with several smoking-related traits.^35,37^ We also identified an association with a deleterious (CADD=18.9)^42^ SNP (rs986391, *p*=3.08E-14, chr. 5) in the *TENM2* gene, recently implicated in smoking initiation, cigarettes per day, and smoking cessation.^13^

Furthermore, we identified variants in *GRM8* (Glutamate Metabotropic Receptor 8; rs2157752, *p*=5.32E-09, chr.7), important for mediating reward-related learning and memory, and in *BDNF* (rs6265, *p*=7.98E-10, chr. 11), a candidate gene in genetic studies of substance use disorders given its role in synaptogenesis and memory. None of the lead SNPs showed evidence of heterogeneity across cohorts, based on the I^2^ index (**Supplementary Figure 4**). Combining these data with UKBB (which uses a less stringent TUD definition, TUD-multi+UKBB) yielded fewer lead SNPs (**Supplementary Table 8**).

### Within-ancestry meta-analyses identify ancestry-specific loci associated with TUD

We conducted within-ancestry meta-analyses of EUR (TUD-EUR) and AA (TUD-AA) using a sample-size weighted fixed effects model, and a GWAS of LA (TUD-LA).

TUD-EUR included 11,422,241 imputed SNPs in a cohort of 163,734 TUD cases and 331,271 controls, which is 8.5 times larger than the total sample size of previous nicotine dependence GWAS.^27^ Observable inflation is attributable to polygenic signal rather than population stratification or other confounding (LDSC intercept 1.049, SE=0.012) and we did not identify evidence of heterogeneity (*I*^2^) across the cohorts (**Supplementary Figure 6**). The TUD-EUR meta-analysis yielded a significant *h^2^_SNP_* estimate of 11.70% (SE=0.005, **Supplementary Table 9**), and identified 74 GWS significant lead SNPs located in 63 independent loci (**Figure 2B**; **Supplementary Table 10**). Fourteen of these loci were ancestry specific in EUR and not GWS in the multi-ancestry GWAS. Among the 63 independent loci, 13 were fine-mapped to a credible set (posterior inclusion probability > 0.50), of which 6 harbored known protein coding genes (*CHRNB2, GALNT10, FAM168A, SPATS2, SYT17, ASIC2*; **Supplementary Table 11**).

Again, combining these data with those of UKBB in a secondary GWAS (TUD-EUR+UKBB) yielded very similar results (e.g., similar *h^2^_SNP_* estimate of 9.30% and *r_g_* estimate of 0.99, SE=0.001; lead SNPs and independent loci presented in **Supplementary Table 12**). Considering the similarity between the primary and secondary GWAS, all downstream analyses used the EUR GWAS for the most stringent TUD definition (TUD-EUR), which excluded the UKBB sample.

The TUD-AA meta-analysis yielded a significant *h^2^_SNP_*estimate of 11.09% (SE=0.014, **Supplementary Table 9**), and 2 independent loci (**Supplementary Table 13**), one on chr. 9 (rs2007153, *p*=1.17E-08) in *DBH*, which is novel for the AA population, and another on chr. 20 (rs6011779, *p*=9.27E-09) in the *CHRNA4* gene, replicating a finding from a prior multi-ancestral (EUR+AA) GWAS of smoking.^27^ Multi-ancestry fine-mapping analyses using PAINTOR corroborated the region in chr. 9, identifying two potential causal variants in this locus (**Supplementary Table 14**). The TUD-LA GWAS yielded a significant *h^2^_SNP_*estimate of 8.14% (SE=0.02, **Supplementary Table 9**) but did not identify any GWS loci (**Figure 2**), presumably due to the smaller sample size.

### Integration with functional genomic data implicates hundreds of novel TUD candidate risk genes

To further our biological interpretation of the TUD-EUR GWAS results and prioritize potential candidate genes and proteins, we performed multiple *in silico* downstream analyses using MAGMA,^41,42^ H-MAGMA,^43^ S-MultiXcan/S-PrediXcan,^44^ TWAS,^45^ and PWAS.^45^

First, we conducted gene-based analyses via MAGMA,^41,42^ which mapped SNP-level associations to 91 significant genes (*p*<2.63E-06), 20 (21.62%) of which replicated genes near or in GWS loci (e.g., *CHRNA3*, *CHRNA4, KDM4A*, *DBH*; **Supplementary Table 15**).

To identify neurobiologically relevant target genes, we incorporated TUD GWAS data with chromatin interaction profiles from human brain tissue using Hi-C coupled MAGMA (H-MAGMA).^43^ These analyses identified 1,017 unique gene-tissue pairs associated with TUD (*p*<9.44E-07), a significant proportion of which showed cell-type (15.63% cortical neurons, 16.42% iPSC-derived neurons, 20.75% midbrain dopaminergic neurons, 14.25% iPSC-derived astrocytes) or developmental stage-specific (15.73% fetal, 17.21% adult) expression (**Supplementary Table 16**).

Using S-MultiXcan to predict the effect of common SNP variation on gene expression in multiple brain tissues, we detected significant associations for 46 genes (**Supplementary Table 17**), with effects dispersed across 13 brain regions (amygdala, anterior cingulate cortex, basal ganglia [nucleus accumbens and putamen], cortex and frontal cortex, cerebellar hemisphere, cerebellum, hypothalamus, spinal cord, substantia nigra). Inspection of region-specific results via S-PrediXcan identified 25 genes that were consistently upregulated (*GPX1*, *PPP6C*, *GMPPB*, *WDR6, QRICH1*, *NICN1*, *ARFRP1*, *METTL21B*, *RNF123*, *CCDC88B*, *HIST1H2BD*, *CCDC71*, *PSMA4*) or downregulated (*CHRNA2, AMT*, *P4HTM*, *NCKIPSD*, *ATP23*, *DALRD3*, *MST1*, *RHCE*, *TSFM*, *RBM6*, *TRIM35*, *PHACTR4*) in more than one brain region (**Supplementary Table 18**).

Next, we assessed differential transcriptomic and proteomic regulation of TUD risk loci in the dorsolateral prefrontal cortex (**DLPFC**) by performing TWAS (mRNA and splicing) and PWAS, respectively. Associations across these three regulatory models identified 50 unique TUD risk genes (32, mRNA expression; 13, splicing expression; 14, proteome expression; **Supplementary Tables 19** and **20**). Colocalization analysis identified four genes and proteins (NT5C2, GPX1, ABHD12, RHCE) associated with TUD via their regulation of brain expression levels and protein abundance (PP4 >0.80, **Supplementary Table 21**, **Supplementary Figure 7**).

Overall, after controlling for multiple comparisons, these analyses identified 461 unique genes with statistical evidence of association with TUD (**Figure 3a**, **Supplementary Table 22**). Of these, 159 genes converged across at least 2 methods, and 2 genes (*GPX1*, *GMPPB*) converged across all six methods and replicated prior GSCAN findings. 110 (23.86%) of the 461 genes identified via these analyses were identified by the GWS loci, and two were novel TUD genes not identified in prior FTND or GSCAN analyses (*PTCHD4*, *THUMPD3*), which prompt novel hypotheses to be tested experimentally.

**Figure 3.**
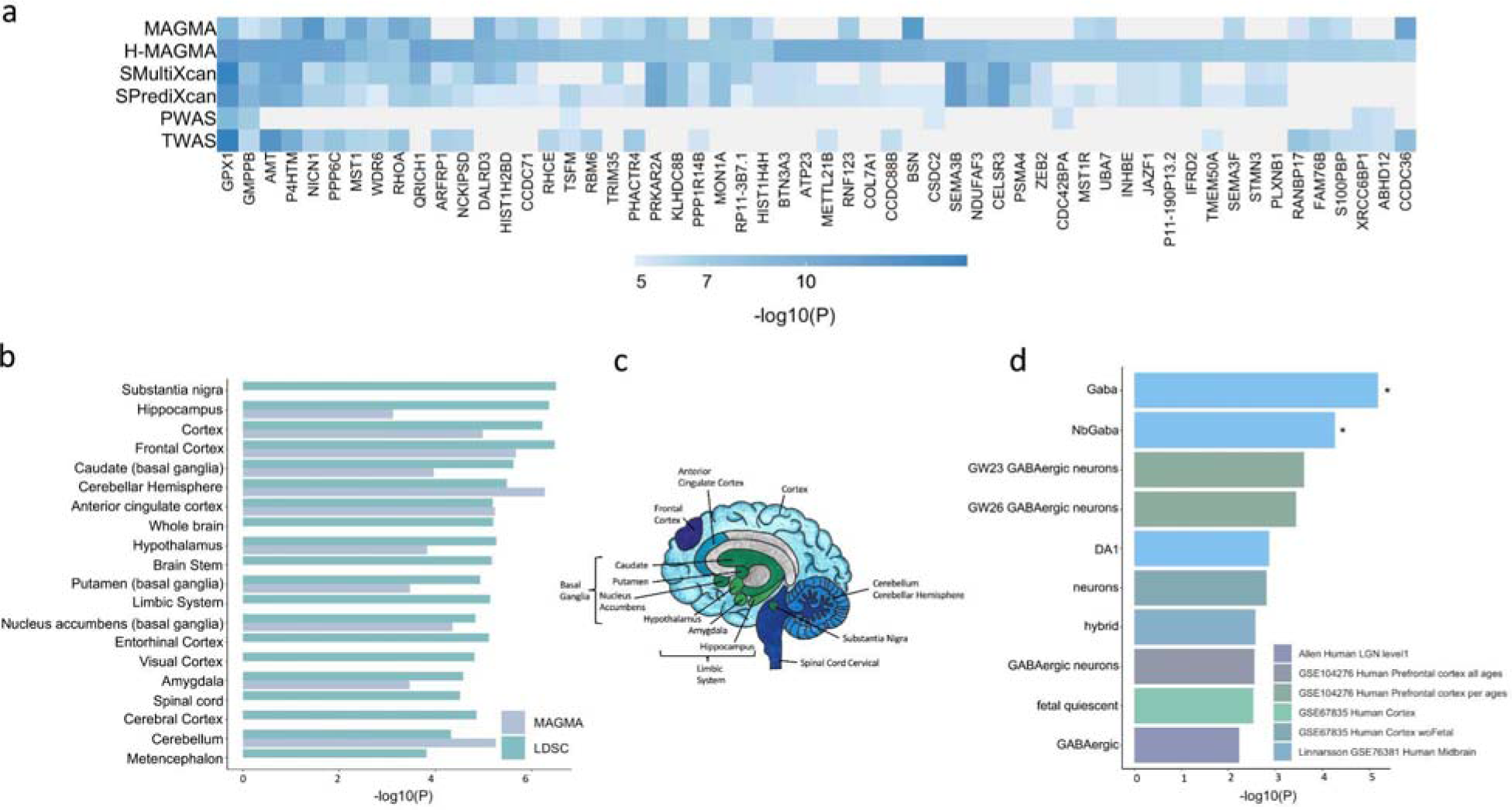
Integration with functional genomic data implicated 461 unique TUD candidate risk genes. (**a**) Of 461 associated genes, 56 converged with at least 3 methods, and were dispersed throughout the chromosomes. (**b**) LDSC (SNP-based) and MAGMA tissue-specific gene expression of TUD risk genes reveals substantial brain enrichment (**Supplementary Tables 25-26**). (**c**) The genetic findings across multiple levels of analysis (LDSC, MAGMA, MultiXcan, BrainXcan) implicated brain regions exhibiting anatomical differences in cases. (**d**) Cell type-specific expression of TUD risk genes. Results from MAGMA property analyses and gene expression using human single-cell RNA-sequencing datasets (**Supplementary Table 28** for full list). After multiple testing correction for all datasets, only genes expressed in GABAergic neurons were associated with TUD (**Supplementary Table 28**).

### Tissue and cell-type analyses of TUD identify enrichment in brain tissue and GABAergic neurons

To identify relevant tissues implicated in TUD, we performed various SNP (LDSC partitioned heritability) and gene-wide (MAGMA) analyses. We performed partitioned heritability in LDSC to evaluate the enrichment of the genome-wide findings in over 50 functional genomic annotations (and across tissues, as described below). In the baseline LDSC model, conserved and regulatory functional annotations were significantly enriched (**Supplementary Figure 8** and **Supplementary Table 23** for full list).

Tissue enrichment analyses in MAGMA use gene expression data from GTEx (v8). In addition to non-brain tissues (i.e., cardiovascular, hematopoietic, adrenal pancreas, and other, *p*<3.37E-05, **Supplementary Table 24**), we detected significant enrichment mostly in the brain (*p*=1.53E-15), spanning multiple brain regions, including the hippocampus, the limbic system, frontal cortex (**Supplementary Tables 25-26**, **Figure 3b-c**), most of which were also implicated in S-MultiXcan (**Supplementary Table 17**). Correlating the effects of SNP variation with brain imaging traits via BrainXcan identified similar results, including significant (*p*<1.92E-04) associations with decreased gray matter volume in the right ventral striatum (**Supplementary Table 27**).

Next, we used FUMA to examine cell-type specific gene expression associated with TUD, leveraging single-cell RNA-sequencing (sc-RNA seq) datasets. After multiple correction testing across datasets, we identified a significant association between TUD risk and cell-type specific gene expression in GABAergic neurons for individual human sc-RNA seq datasets (Linnarsson, midbrain, gaba: *p*<5.03E-03; nbGaba: *p*<4.29E-02; **Figure 3d**; **Supplementary Table 28**). These results did not survive conditional analyses within and across datasets.

### Implications for TUD biology based on gene-set and pathway analyses

We used MAGMA^41,42^ to conduct a gene-wise TUD analysis and to test for enrichment of pathways curated from multiple sources. After correcting for multiple comparisons, 13 related pathways and biological processes were significantly enriched for genes associated with TUD (*p*<2.65E-06; **Supplementary Table 29**). Associations implicated fundamental processes related to nicotine response (e.g., high calcium and sodium permeable nicotinic acetylcholine receptors, *p*=6.03E-15; behavioral response to nicotine, *p*=5.81E-13), regulation of postsynaptic nicotinic acetylcholine receptors (*p*=1.32E-10), and nicotine effect on dopaminergic neurons (*p*=1.87E-06), among others.

### Drug Repurposing

Linking transcriptome-wide patterns to perturbagens that pass the blood-brain barrier from the Library of Integrated Network-Based Cellular Signatures (LINCS)^36^ database identified 235 medications approved by the U. S. Food and Drug Administration (**Supplementary Table 30**). Of the 235 identified medications, 20 targeted at least one mapped/independent gene from our GWAS (**Figure 4**). The medications that significantly reversed (Bonferroni *p*<6.03E-05) the transcriptional profile associated with TUD included varenicline (a well-known therapeutic for smoking cessation), sodium channel blockers (e.g., amiloride), and compounds that are used to treat conditions that commonly co-occur with TUD, such as antipsychotics (e.g., clozapine), dopaminergic agents (e.g., ropinirole), opioids (e.g., nalbuphine), and antidepressants (e.g., amoxapine), among others (**Supplementary Table 30**).

**Figure 4.**
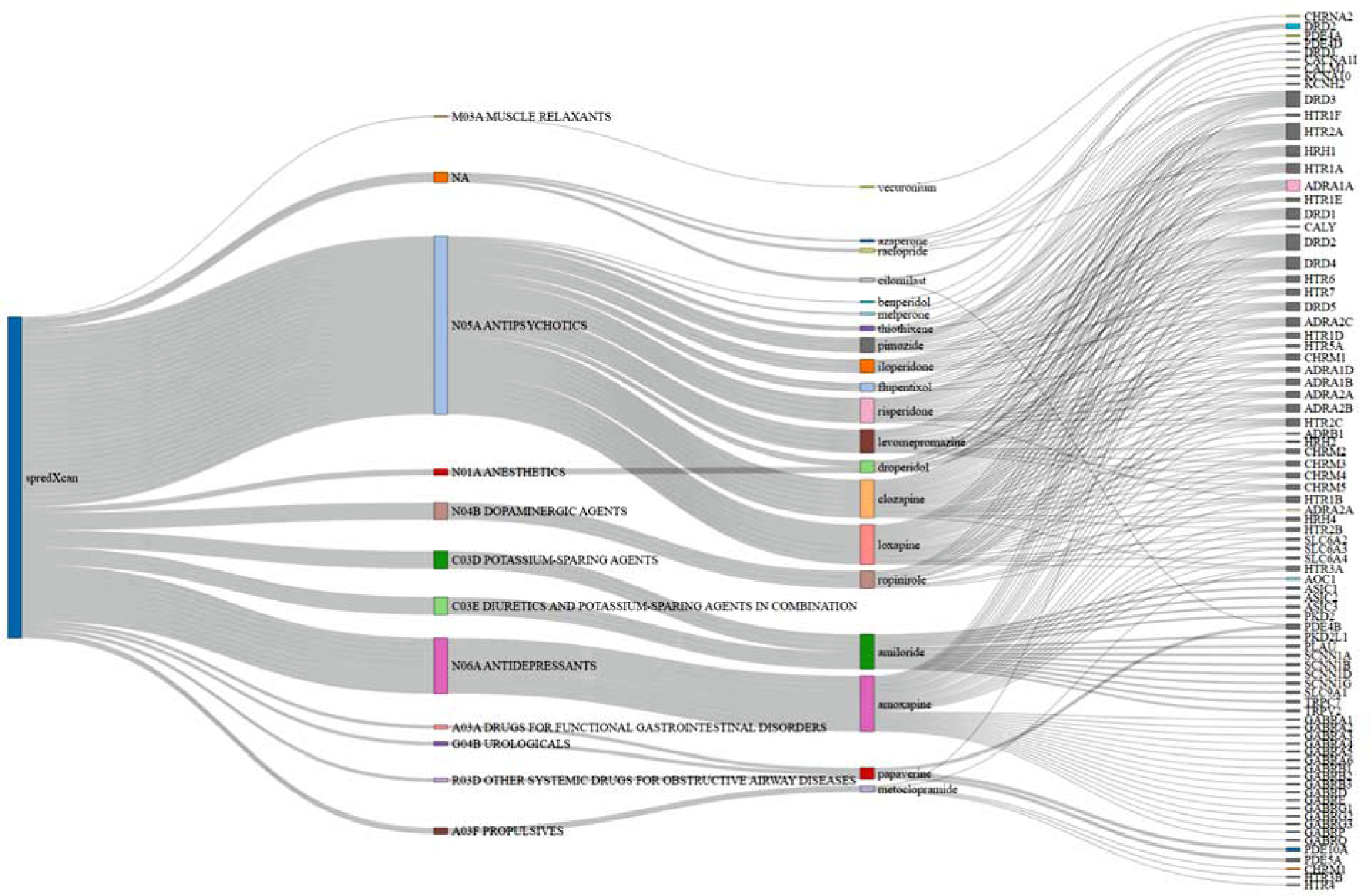
Sankey Diagram showing drug repurposing results from S-PrediXcan brain tissues. 20 medications/perturbagens grouped by ATC category membership from the Library of Integrated Network-Based Cellular Signatures (**LINCS**)^36^ database. ATC categories connected to perturbagen edges represent corresponding ATC category membership. Perturbagens connected to gene target edges are associated with the reversal of the TUD transcriptomic profile from S-PrediXcan brain tissue results. Only medications that targeted at least one mapped/independent gene from our GWAS are plotted.

An additional drug repositioning analysis using DRUGSETS identified three significant (Bonferroni *p*<6.80E-05) medications: varenicline, cytisine, and galantamine (**Supplementary Table 30**).

### Genetic correlation with other traits

We estimated pairwise *r_g_* with TUD for 113 published phenotypes using LDSC.^46^ TUD showed FDR-significant correlations *r_g_* with 76 traits (**Figure 5b**; **Supplementary Table 31**). As expected, the strongest positive correlations were with smoking-related traits (e.g., age of smoking initiation *r_g_*=-0.59, SE=0.03; smoking initiation *r_g_*=0.81, SE=0.02; cigarettes per day *r_g_*=0.44, SE=0.03; smoking cessation *r_g_*=0.66, SE=0.02; FTND *r_g_*=0.63, SE=0.06; **Figure 5a**) and other substance use traits (e.g., cannabis use disorder *r_g_*=0.64, SE=0.04; drinks per week *r_g_*=0.36, SE=0.02; opioid use disorder (**OUD**) *r_g_*=0.47, SE=0.04). TUD clustered with addiction traits rather than consumption phenotypes (**Supplementary Figure 9**).

**Figure 5.**
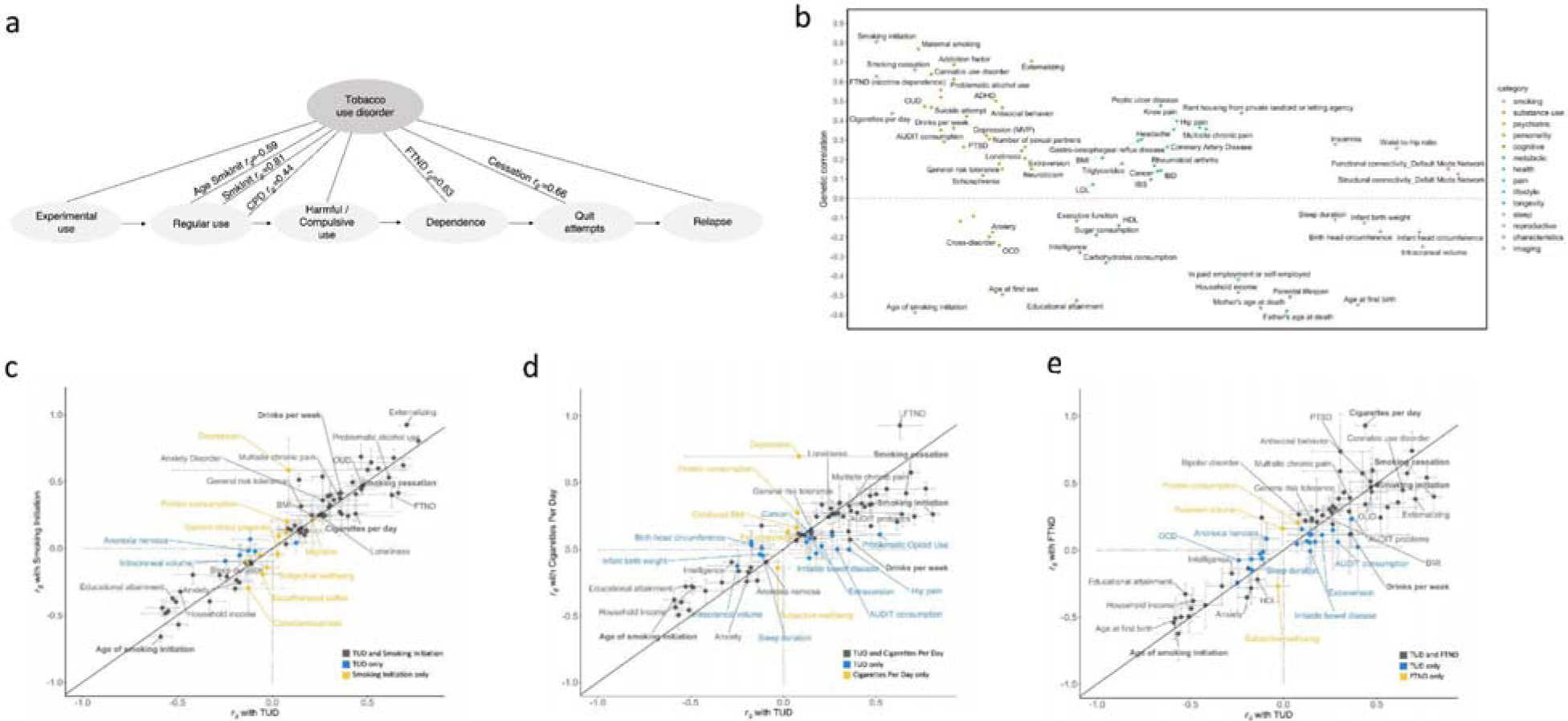
FDR-significant genetic correlations between TUD-EUR and 113 complex traits, including smoking and related phenotypes (b). (**a**) TUD consists of multiple components, progressing from experimental use to regular use, compulsive use, cessation, and relapse. Therefore, high genetic correlations (*r_g_*) are to be expected between the age of smoking initiation (AgeSmkInit), smoking initiation (SmkInit), cigarettes per day (CPD), smoking cessation (SmkCess)^13^, nicotine dependence measured using the Fagerström Test for Nicotine Dependence (FTND)^27^, and tobacco use disorder (see **Supplementary Table 31** for full results). (**b**) Genetic correlations with an extended list of traits from publicly available GWAS. Traits with positive *r_g_* values are plotted above the line; traits with negative *r_g_* values below the line. All *r_g_*s are significant using a 5% FDR correction for multiple testing. (**c-e**) Systematic comparison of significant genetic correlation estimates between TUD and SmkInit (c), CPD (d) and FTND (e) reveal overlapping (black dots) and trait-specific (blue and yellow dots) relations between TUD and these other smoking phenotypes. *r_g_* estimates were generally higher for TUD than CPD - even with a smaller sample size (TUD, N=495,005; CPD, N=784,353) - and FTND. On the contrary, *r_g_*’s were generally smaller for TUD than SmkInit, possibly because of the larger sample for SmkInit (N=3,383,199) than TUD. Overall, these results indicate that these smoking behaviors, including SmkInit, CPD, FTND, and TUD, represent both unique and interrelated polygenic influences, which are complementary to those associated with other complex behaviors and disorders at the genetic level.

TUD was also genetically associated with 59 other psychiatric and medical conditions (**Figure 5b**, **Supplementary Table 31**). There were significant positive *r_g_* with psychiatric traits (e.g., externalizing *r_g_*=0.71, SE=0.02; ADHD *r_g_*=0.50, SE=0.03; posttraumatic stress disorder *r_g_*=0.31, SE=0.08) and risky behavioral traits, including younger age of first sex (*r_g_*=-0.50, SE=0.03). We also found positive *r_g_* with health outcomes (e.g., coronary artery disease *r_g_*=0.26, SE=0.03; waist-to-hip ratio *r_g_*=0.26, SE=0.02; multisite chronic pain *r_g_*=0.36, SE=0.03) and several social determinants of health, such as the Townsend deprivation index (*r_g_*=0.61, SE=0.07). There were negative *r_g_* with socioeconomic variables, including educational attainment (*r_g_*=-0.53, SE=0.02) and household income (*r_g_*=-0.49, SE=0.03) and with intelligence (*r_g_*=-0.28, SE=0.02). Conditioning on alcohol, cannabis, or opioid use disorder did not substantially modify the magnitude or direction of these associations (**Supplementary Table 32**). Virtually all *r_g_* estimates for other phenotypes were greater with TUD than cigarettes per day (**Figure 5c**) and FTND (**Figure 5d**), but not smoking initiation (**Figure 5e**).

Among AA samples, there were significant *r_g_* with smoking trajectories and other substance use traits (OUD *r_g_*=0.44, SE=0.11; maximum habitual alcohol consumption *r_g_*=0.77, SE=0.19). Nominal associations (*p*<0.05) were observed for smoking initiation (*r_g_*=0.35, SE=0.13), depression (*r_g_*=0.45, SE=0.22) and type 2 diabetes (*r_g_*=-0.23, SE=0.09; **Supplementary Table 33**).

### Phenome-wide association analyses

To further explore pleiotropic effects, we performed a series of phenome-wide association studies (**PheWAS**) of TUD polygenic scores (**PGS**) in other EHR and clinical cohorts of adults, and a young population-based cohort. We performed these analyses within ancestries.

#### EHR cohorts

We conducted PheWAS with EHR data to test the association between polygenic risk for TUD and liability for thousands of other medical conditions, including TUD, in another independent site, Mayo Clinic. As expected, TUD PGS was strongly associated with TUD (*p*=1.90E-145, **Supplementary Table 34**, **Figure 6a**), explaining 7.3% of the (Nagelkerke’s *R*^2^) variance. Additional significant (*p*<3.24-05) associations included 4 traits in the substance use disorders domain (e.g., alcohol-related disorders, OR=1.33, *p*=6.30E-26), 10 psychiatric conditions (e.g., depression, OR=1.09, *p*=4.31E-11), and medical conditions strongly associated with TUD (e.g., chronic airway obstruction, OR=1.25, *p*=1.60E-32). Most of these associations remained significant after accounting for TUD diagnosis (**Supplementary Table 34**). We also noted associations across multiple other medical categories, including endocrine/metabolic (e.g., morbid obesity, OR=1.12, *p*=3.53E-13; type 2 diabetes, OR=1.09, *p*=1.48E-09), digestive (e.g., diseases of esophagus, OR=1.07, *p*=1.47E-10), circulatory (e.g., ischemic heart disease, OR=1.09, *p*=1.56E-11) and neurologic (e.g., pain, OR=1.07, *p*=4.33E-08), among others (**Supplementary Table 34**). Compared to FTND PGS, TUD PGS were more strongly associated across virtually all domains, including TUD (**Figure 6a**). After conditioning on PGS for other smoking variables (CPD, SmkInit, FTND), TUD PGS was still significantly associated with TUD and 14 other mental and medical traits (**Supplementary Table 34**). We repeated the TUD PGS analyses in a BioVU cohort of AA individuals using the TUD-AA meta-analysis results. As expected, TUD was the strongest and most significant (OR=1.20, *p*=2.81E-06) association (**Supplementary Table 35**).

**Figure 6.**
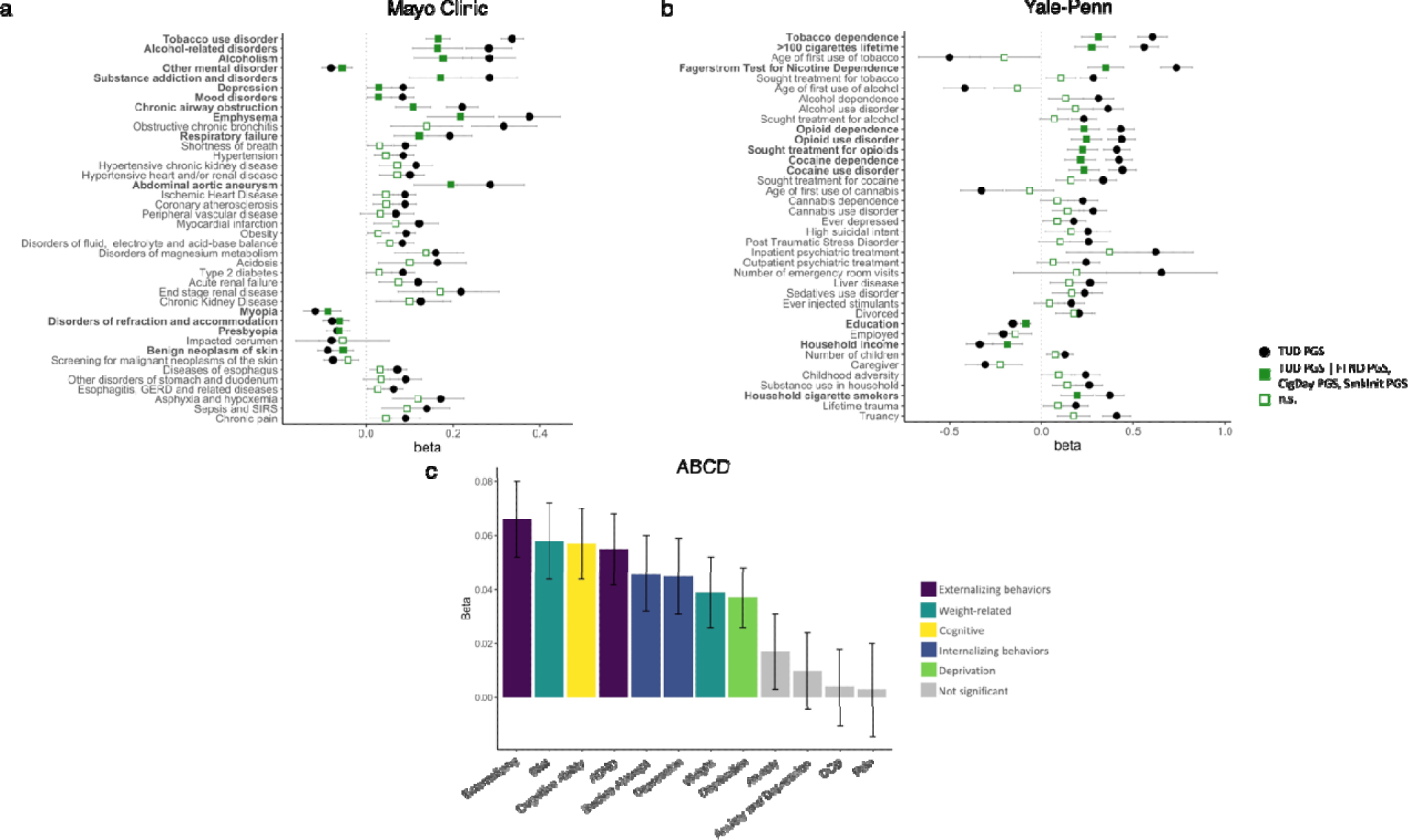
TUD PGS PheWAS in the (a) Mayo Clinic, (b) Yale-Penn, and (c) ABCD European cohorts. Only selected Bonferroni-significant traits are shown. In (a) and (b), association of TUD PGS (in black) is conditioned on PGS for FTND, CPD, and SmkInit (in green). The exact values for each association and extended lists of traits can be found in **Supplementary Tables 34, 36 and 38**.

#### Yale-Penn sampl

We next extended the analyses to a deeply characterized sample recruited for genetic studies of substance use disorders: the Yale-Penn sample.^47^ We examined the association between PGS for TUD and hundreds of other traits derived from a comprehensive psychiatric interview, the Semi-Structured Assessment for Drug Dependence and Alcoholism (**SSADDA**). TUD-EUR and TUD-AA PGS were strongly associated with nicotine dependence as defined via a Diagnostic and Statistical Manual of Mental Disorders (**DSM**) diagnosis in both the EUR (OR=1.83, *p*=3.51E-49; **Figure 6b**; **Supplementary Table 36**) and AA cohorts (OR=1.13, *p*=7.13E-04), respectively, although the latter association did not survive multiple testing correction (**Supplementary Table 37**). In the EUR cohort, we also noted significant associations between TUD-EUR PGS and 224 other phenotypes, including 163 in the substance-related domain (44 opioid-related, 31 cocaine-related, 25 alcohol-related, 23 tobacco-related, 14 sedative-related, 13 cannabis-related, 10 other, 2 stimulant-related), and 50 in other domains (13 medical, 33 psychiatric [9 PTSD, 11 depression, 7 antisocial personality, 3 suicide, 2 ADHD, and 2 conduct disorder], 9 environmental, and 6 demographic phenotypes. Again, compared to FTND PGS, TUD-EUR PGS was more strongly associated across virtually all domains, including nicotine dependence (Nagelkerke’s *R*^2^=0.101 vs 0.062; **Supplementary Table 36**). After conditioning on PGS for other smoking variables (CPD, SmkInit, FTND), TUD PGS was still significantly associated with 11 smoking-related traits and 50 other mental and medical conditions (**Supplementary Table 36**), again emphasizing the value of collecting information on later stages of vulnerability or more severe phenotypes, such as TUD.

#### Adolescent Brain Cognitive Development (ABCD) cohort

Lastly, we extended our polygenic analyses to a drug naïve developmental sample (9-11 years of age at recruitment; analytic N=62 to 5,556). We concentrated on 12 traits that showed significant genetic correlations in the adult samples (**Supplementary Table 38**, **Figure 6c**). Although tobacco exposure was uncommon in this pediatric population (2.30% prevalence), externalizing behaviors, which emerge in childhood and are strong correlates of substance use, were available. After correcting for multiple testing, TUD PGS was significantly (*p*<4.00E-03) associated with externalizing behaviors (i.e., Child Behavior Check List [**CBCL**] externalizing scores, β=0.07, *p*=1.21E-06; CBCL ADHD scores, β=0.06, *p*=4.97E-05), as well as internalizing (i.e., suicide attempt, β=0.05, *p*=1.52E-03, CBCL depression scores, β=0.05, *p*=1.11E-03), cognitive ability (β=0.06, *p*=8.35E-06), neighborhood deprivation (β=0.04, *p*=1.05E-03), and weight-related phenotypes (i.e., BMI, β=0.06, *p*=1.61E-05; weight, β=0.04, *p*=2.77E-03). Notably, these children were not chronically exposed to tobacco; therefore, we would speculate that these associations are not a consequence of smoking but rather may underlie overlapping genetic architectures among the traits studied that predate use of tobacco.

### Causal relationships with TUD and bi-directional effects of TUD with other traits

We used MR analyses to test directional causal relationships between significantly genetically correlated traits (N=31) and TUD among EURs only due to the small sample size and limited statistical power in other populations (**Supplementary Table 39**). There was a positive causal effect of TUD on cross-disorder. Seven traits showed significant causal effects on TUD. Specifically, we observed a negative causal effect of education attainment, and a positive causal effect of drinks per week, depression, BMI, externalizing, opioid prescriptions, and opioid use disorder on TUD.

## Discussion

Uncovering the genetic underpinnings of individual differences in TUD liability can advance diagnosis, prevention, and treatment efforts for a disorder of enormous public health significance. GWAS have uncovered multiple associations with tobacco use, but findings for tobacco dependence or disorder have been limited due to the difficulty of characterizing large numbers of individuals using a gold-standard research or clinical diagnosis. Here we present the first multi-ancestry GWAS of TUD using data from EHR, as a complementary strategy for ascertainment. EHR-biobanks are the result of years of work recruiting, consenting, and genotyping individuals. As a result, researchers can now conduct studies such as the one reported here, gathering data for 898,680 individuals in less than 4 months, to identify novel biology for disorders. The number of GWAS signals, enrichment in relevant pathways and tissues, and genetic overlap with nicotine-related traits provide proof of principle that EHR can serve as a complementary tool to study TUD genetics.

Our findings demonstrate that TUD, as defined via EHR, was genetically correlated with traits derived from traditionally ascertained cohorts, including nicotine dependence via FTND and smoking cessation, providing clear evidence that the signal captured by TUD phecodes is valid. Of note, the genetic correlation between TUD and cigarettes smoked per day (**CPD**) was relatively modest (*r_g_*=0.44), suggesting that the genetic architectures of consumption and misuse are only partially overlapping, consistent with prior GWAS of alcohol and cannabis use and misuse (e.g., ^23,26,48^). This contrasts with earlier observations for FTND and CPD, for which the genetic correlation was almost at unity (*r_g_*=0.95).^27^ This shows that TUD captures features beyond the frequency of smoking or severity of nicotine dependence. Although FTND and TUD were more strongly genetically correlated (*r_g_*=0.63), in general, we observed that TUD PGS was more predictive of DSM-defined tobacco dependence and a plethora of comorbid traits in the Yale-Penn sample, than FTND PGS. The only exception was for smoke after waking, which was more strongly associated with FTND PGS, likely because time-to-first cigarette is one of the FTND items. TUD was highly correlated (*r_g_*=0.81) with regular cigarette use (i.e., smoking at least 100 cigarettes in a lifetime, previously referred to as “smoking initiation”)^13^, which is expected as nicotine is a highly addictive substance, with 85% of smokers meeting criteria for TUD.^1,2^ However, our polygenic findings demonstrate that TUD explains additional variance above and beyond that accounted for by other smoking traits (smoking initiation, CPD, FTND). This emphasizes the need to measure the full spectrum of addiction liability,^49^ from regular use to more severe phenotypes, such as TUD, to account for the distinct biological factors relevant at each stage.

Common SNPs were able to account for a fraction (12%) of the overall heritability of TUD (40-60%) as determined by prior family and twin studies.^9,11^ The multi-ancestral meta-analysis identified 88 independent loci, 18 times the number previously reported for nicotine dependence.^27^ These include corroborative support for the involvement of nicotinic acetylcholine receptor genes (*CHRNA5-A3-B4*, *CHRNB2*, *CHRNA2*, *CHRNA4*), which have been consistently associated with smoking behaviors,^20^ particularly in studies of self-reported CPD.^13^ Other variants identified are in genes that modulate dopaminergic and glutamatergic neurotransmission, compromising reward-based learning and facilitating drug-seeking behavior, and in *BDNF*, which is involved in memory consolidation processes,^51^ and a well-studied candidate gene in addiction.^52^ These and other candidates supported by TUD (e.g., *PDE4B*) were genetically correlated with other addiction phenotypes,^36^ emphasizing the shared neurobiological mechanisms of addiction.

Downstream analyses prioritized genes and drug candidates that could be used for follow-up mechanistic studies in model organisms. Specifically, we identified “core” genes that could be “pleiotropic hotspots” associated with multiple traits. One was glutathione peroxidase-1 (*GPX1*), which is involved in oxidative stress. Intriguingly, it has been reported that glutathione peroxidase-1 protects against lung inflammation induced by smoking in mice, and agents that mimic this action (e.g., ebselen), which restore GPX1 activity in situations of extreme oxidative stress, can protect from lung inflammation induced by smoking.^53^ Another was *GMPPB*, which has been associated with accelerated lung aging and e-cigarette smoking.^54^ *NT5C2* is involved in maintaining cellular nucleotide balance, and was associated with schizophrenia^55^ and smoking behaviors in an exome-wide association study.^56^ These genes showed a consistent association based on colocalization analyses (here and previously^57^), suggesting that they could confer TUD risk by modulating regulated gene expression and protein abundance in the brain.

The enrichment of TUD in brain tissues further supports TUD as a brain disorder, long supported by neuroscience and more recently by genetics.^58^ We provide suggestive evidence for the involvement of the cerebellum in TUD, along with other regions that have long been studied in relation to addiction such as the fronto-striatal loop, hippocampus, and amygdala.^59^

Genetic correlations revealed substantial levels of pleiotropy with traits that often co-occur with TUD, including other substance use and psychiatric disorders. These associations were particularly evident in the Yale-Penn sample,^47^ which has comprehensive phenotypic data for substance use disorders. In adult patients from the Mayo Clinic, we replicated the associations with substance and other psychiatric disorders, extending them to medical disorders, such as HIV, heart disease, and pain, some of which (e.g., respiratory conditions) likely reflect chronic smoking. The positive associations between genetic liability for TUD and other outcomes, such as BMI and other internalizing/externalizing problems in tobacco-naive children (ABCD), may also reflect true biological relationships. Although we are far from untangling this complex web of genetic and non-genetic correlations, the extensive phenotypic spectrum associated with TUD is undeniable.

Currently, developing new therapeutics for TUD is viewed as risky because of a lack of high-quality targets, historically low success rates, and unintended side effects. Although genes identified in our GWAS, including *CHRNA5*, *CHRNA4*, and *CHRNB2*, might moderate the effect of varenicline, a smoking cessation treatment that operates as a partial agonist at the nicotine acetylcholine a2b4 receptor,^60^ varenicline (along with other medications such as nicotine replacement therapies) has limited efficacy or adverse effects.^61,62^ In a proof-of-principle study, So et al.^63^ identified several repurposing candidates for treating psychiatric disorders by connecting imputed transcriptomic profiles from GWAS data to drug-induced gene expression profiles. Using this approach, we identified hundreds of potential drug candidates predicted to significantly reverse the TUD transcriptomic profile. These included norepinephrine reuptake inhibitors (e.g., amoxapine) and antipsychotics (e.g., clozapine), pointing to convergent molecular mechanisms between TUD and other psychiatric disorders that are the usual target of these agents, replicating prior observations.^64,65^ The potential therapeutic utility of anti-inflammatory and blood glucose lowering medications was also suggested by our analyses, in addition to an anti-Parkinson medication known to interact with dopaminergic activity (i.e., biperiden) and one that acts both as an antagonist of acetylcholinesterase and an agonist of nicotinic receptors (i.e., galantamine), as shown in recent independent studies.^65,66^ Although, to date, no repurposed drugs have been developed for treating SUDs based on GWAS data, this is an important potential path forward, particularly for SUDs, where few effective pharmacotherapies are available.

Future research may address some of the limitations of our study. Prior work has demonstrated that ICD codes have a low sensitivity for current tobacco use, but may have a reasonable specificity for this common behavior.^67^ Our results appeared to be robust to moderate levels of misclassification, particularly in controls, as detected by the pairing with self-reported questionnaire data. Our results also appeared to be robust to moderate levels of cross-cohort heterogeneity, including potential differences in diagnostic practices and different levels of misdiagnosis of control populations across sites. Although studies that systematically evaluate the effect of removing potentially misclassified individuals are needed, we chose not to remove them in this study because not all individuals had concomitant survey data available. This questionnaire data, along with other forms of EHR data (e.g., clinical notes), may help capture additional phenotypes, including the response to treatment or the ability to successfully quit smoking without formal treatment. We have highlighted potential differences of traits ascertained by ICD codes as a limitation of our study. *r_g_* results revealed high levels of association between TUD and hundreds of other traits. However, the extent to which TUD shares biological underpinnings with other traits and diseases may also be influenced by potential misdiagnosis, ascertainment and cross-trait assortative mating, among many other factors.^68^ Longitudinal data from EHR, with data collection spanning the period prior to and following the onset of substance use and SUD, are particularly valuable for studying the timing of onset, within-person change, and application of time-varying effects, which will help to differentiate causation from correlational findings. The advent of single-cell transcriptomics, larger QTL databases in more specific cell types, and the inclusion of more ancestrally diverse samples will improve the interpretability of associated loci. Although we have included diverse cohorts, our study lacked many major ancestral groups such as East Asians and South Asians. Furthermore, other forms of genetic variation, such as rare single variants^69^ or structural polymorphisms^70^ are likely to account for much of the “missing heritability” in genetic risk for TUD. Lastly, tobacco use can be greatly affected by environmental factors,^12^ such as cultural context, public health policies and characteristics related to socioeconomic status.^71^ Together with the existing body of literature,^72–75^ the strong genetic correlations between TUD and environmental influences, such as Townsend deprivation index, educational attainment and prenatal smoking, underscore the importance of considering environmental moderators in understanding the complex etiology of TUD. There is a great need in the field, therefore, to systematically assess sociocultural factors in healthcare settings.^76^

In sum, this work demonstrates that EHR is a viable and cost-efficient complementary approach to rigorous clinical ascertainment for genetic studies of TUD, similar to other SUD traits. At various levels of analysis, this study identifies and prioritizes previously unidentified genes of potential interest. TUD shares biological processes common to many SUDs and is highly correlated with many psychiatric and medical disorders. We anticipate that these results can be combined with prior smoking GWAS in larger multivariate analyses to elucidate the full spectrum of smoking behaviors and accelerate gene discovery for TUD.

## Methods

### Smoking phenotypes and cohorts

We defined cases as patients who received at least two TUD ICD-9 or -10 codes (corresponding to the phecode definition) in their medical records, and controls as patients who had no TUD diagnosis codes (**Supplementary Table 2**). In UKBB only, cases were defined as having 1 ICD-10 code for TUD, and controls had none.^41^ Additionally, we required controls to be 18 years of age or older at time of analysis (04/2022). Patients younger than 18 years were excluded because they may not yet have reached the age of TUD diagnosis. We examined the sensitivity of our TUD phenotyping using the patients’ self-reported tobacco use via survey data when available (**Supplementary Table 3,** list of smoking traits).

Our data sources included registries from five health systems linked to biobanks: Vanderbilt University Medical Center’s (**VUMC**) biobank (**BioVU**), Mass General Brigham Biobank (**MGBB**), Penn Medicine Biobank (**PMBB**), Million Veteran Program (**MVP**), and UK Biobank (**UKBB**). There were 46,905 (EUR) patients from VUMC, 22,268 (EUR) patients from MGBB, 39,087 patients from PMBB (28,999 EUR and 10,088 AA), 545,530 patients from MVP (396,833 EUR, 104,332 AA, 44,365 LA), and 244,890 participants from UKBB. Details of each registry, including demographics and data sources, are listed in **Supplementary Table 2**.

### Genotyping, imputation, and GWAS

For all cohorts, the initial GWAS analyses were conducted within genetic ancestral groups. Genetic ancestral groups were determined for BioVU, MGBB and PMBB based on principal component analysis (**PCA**) and comparison to known ancestries in the 1000 Genomes Project Phase 3^77^ reference panel. In MVP, genetic ancestral groups were determined by harmonizing genetic ancestry and self-identified ancestry (HARE),^78^ which also defines genetic ancestry based on the 1000 Genomes reference panel. Further details for each site are included below. GWAS analyses were performed within each ancestral group using SAIGE version 0.44.6.5^79^ or PLINK 2.0^80^ and a logistic regression. For the BioVU, MGBB, and UKBB cohorts, there were GWAS for only the European ancestral group (**Supplementary Material**). In PMBB, we conducted additional GWAS of the African ancestral group sample, and in MVP we performed additional GWAS of the African American ancestral group sample and the Latin American ancestral group sample. Each of the univariate GWAS covaried for 10 genetic ancestry principal components (**PC**), age, sex, number of ICD codes and length of record. The summary statistics for TUD in UKBB were downloaded from the GWAS atlas (https://atlas.ctglab.nl/traitDB/3439).

#### BioVU

We used de-identified clinical data from individuals in BioVU. Genotype data were generated using the Illumina Multi-Ethnic Genotype Array (MEGAEX) for 72,824 individuals. Details on the quality control process have been described elsewhere.^81^ Genotypes were filtered for SNP (<0.95) and individual (<0.98) call rates, sex discrepancies, and excessive heterozygosity (|Fhet|>0.2).^82^ The sample was then filtered for cryptic relatedness by removing one individual of each pair for which pihat>0.2. PCA using FlashPCA2 combined with CEU, YRI and CHB reference sets from the 1000 Genomes Project Phase 3^77^ was implemented to determine European ancestry. We confirmed the absence of genotyping batch effects. We imputed genotypes using the Michigan Imputation Server with the reference panel from the Haplotype Reference Consortium. SNPs were filtered for imputation quality (*R*^2^ >0.3 or INFO >0.95) and converted to hard calls. We restricted the analyses to autosomal SNPs with minor allele frequency (**MAF**)<0.01. We removed SNPs that differed by >10% from the 1000 Genomes Project phase 3 CEU set^77^ and those with a Hardy Weinberg Equilibrium (**HWE**) *p*<1.00E-10. The resulting data set contained hard-called SNP information for 9,386,383 SNPs in 72,824 individuals of European Ancestry. Controls were also required to have 3 or more years of medical history with VUMC. These procedures resulted in a total sample of 7,167 cases and 39,738 controls in BioVU. The project was approved by the VUMC Institutional Review Board (**IRB**, #160302, #172020, #190418).

#### MGBB

MGBB samples were genotyped using the Illumina Multi-Ethnic Global array with hg19 coordinates. Variant-level quality control filters were applied to remove variants with a call rate <0.98, and those that were duplicated across batches, monomorphic, not confidently mapped to a genomic location, or associated with genotyping batch. Sample-level quality control filters were applied to remove individuals with a call rate <0.98, excessive autosomal heterozygosity (±3 standard deviations from the mean), or discrepant self-reported and genetically inferred sex. PCs of ancestry were calculated using the 1000 Genomes Phase 3 dataset as a reference panel. The Michigan Imputation Server was then used to impute missing genotypes with the Haplotype Reference Consortium dataset serving as the reference panel. Imputed genotype dosages were converted to hard-call format and subjected to further quality control, where SNPs were removed if INFO score <0.8, MAF <0.001, HWE *p*<1.00E-10, or missingness (variant call rate <0.98). Only unrelated individuals (pi-hat <0.2) of European ancestry were included in the present study. These procedures yielded a final analytic sample of 6,708 cases and 15,560 in the MGBB. The project was approved by the MGBB IRB (#2018P002642).

#### PMBB

PMBB samples were genotyped by the GSA genotyping array. Quality control removed SNPs with marker call rate <95% and sample call rate <90%, and individuals with sex discrepancies. Genotype phasing and imputation was performed on the TOPMed Imputation server.^83^ The phasing was done using EAGLE (v2.4.1)^30^ and imputation was performed using MINIMAC software.^83^ IBD analysis was used to check for relatedness among imputed samples using PLINK 1.9. We randomly removed one individual from each pair of related individuals (pi-hat <0.25). SNPs with an INFO score <0.3, MAF <0.01, a genotype call rate <0.95 or an HWE *p*<1.00E-6 were removed. To estimate genetic ancestry, PCs were calculated based on common SNPs between PMBB and the 1000 Genomes Project phase3^77^ using the *smartpca* module of the Eigensoft package.^84^ Participants were assigned to an ancestry based on the distance of 10 PCs from the 1000 Genomes reference populations. The resulting dataset included 10,088 AA individuals (cases=1,722) and 28,999 EUR individuals (cases=3,088). The project was approved under IRB protocol #813913.

#### MVP

MVP samples were genotyped using the Affymetrix Axiom Biobank Array. Samples were removed if they had extreme heterozygosity, call rate <98.5%, sex mismatch, or >7 relatives. SNPs were removed if they had call rate <0.98 or a HWE threshold of *p*<1.00E-06. Genotype phasing and imputation was performed using SHAPEIT4 (v.4.1.3)^85^ and Minimac4 software^83^, respectively. Biallelic and non-biallelic SNPs were imputed using the African Genome Resources and 1000 Genomes reference panels.^77^ Ancestry was defined for three mutually exclusive ancestral groups (European, African American, and Hispanic American) utilizing a previously defined approach that harmonizes genetic ancestry and self-identified ancestry (**HARE**).^78^ SNPs with imputation quality (**INFO**) score□<0.7, MAF (AA <□0.005; EUR□<□0.001; HIS□<□0.01), genotype call rate <0.95, and HWE *p*<1.00E-06 were removed. We also excluded one individual from each pair of related individuals (kinship >0.08, N=31,010). The final sample comprised 104,332 AA individuals (cases=43,743), 396,833 EUR individuals (cases=146,771) and 44,365 LA indivisuals (cases=12,277). The Central VA IRB and site-specific IRBs approved the MVP study.

### SNP-heritability (*h*^2^*_SNP_*)

We estimated *h*^2^*_SNP_*based on the liability-scale (population prevalence estimates of 0.125) for common SNPs mapped to HapMap3^86^ using LDSC.^46^ For AA and LA, we created in-sample LD scores derived from the MVP genotype data using cov-LDSC.^87^

### Meta-analyses and independent variants

Meta-analyses were conducted using a sample-size-weighted method in METAL,^88^ assuming shared risk effects across ancestries. Effective sample sizes (**N_Eff**), calculated using the formula: 4/[1/n_case + 1/n_control], were used to compensate for the imbalance in the ratio of cases to controls. N_Eff were used in all meta-analyses and all downstream analyses.

We conducted five meta-analyses of TUD GWAS summary statistics across the following datasets: 1) within-ancestry meta-analysis for EUR samples in BioVU, MGBB, PMBB, MVP, and an additional meta-analysis including UKBB, 2) within-ancestry meta-analysis for AA in MVP and Penn, and 3) multi-ancestry meta-analysis across EUR (BioVU, MGBB, PMBB, MVP), AA (PMBB, MVP), and HA (MVP) datasets, and an additional meta-analysis including UKBB. Inflation of test statistics due to polygenicity or cryptic relatedness was assessed using the LDSC attenuation ratio [(LDSC intercept - 1)/(mean of association chi-square statistics - 1)]. Resulting genome-wide significant (**GWS**) loci were defined as those with *p*<5.00E-08 with LD *r*^2^>0.1, within a 1MB window, based on the structure of the Haplotype Reference Consortium (HRC) multi-ancestry reference panel for the multi-ancestry meta-analysis, or the HRC ancestry-appropriate reference panel otherwise. GWS loci were examined for heterogeneity across cohorts via the *I*^2^ inconsistency metric.

To identify TUD risk loci and lead SNPs, we performed LD clumping in FUMA^41^ using a range of 3□Mb, *r*^2^□>0.1, and the respective ancestry 1000 Genome reference panel.^77^ Genomic risk loci that were located <1Mb apart were incorporated into a single locus. For loci that harbored multiple variants, we used COJO in GCTA^89^ to define independent variants by conditioning them on the most significant variant within each locus. Following conditioning, significant variants (*p*<5.00E-08) were considered independent.

We determined credible variants among the independent variants by merging risk variants within 1Mb of the lead variant and fine-mapped the resulting region with 95% credible sets using FINEMAP.^90^ Posterior inclusion probability ranges from 0 to 1 with values closer to 1 indicating greater causal probability. We implicated a putative causal variant if it accounted for > 50% of the posterior probability in the 95% credible set.

### Multi-ancestry fine-mapping analyses

We used PAINTOR v3.1^91^ to perform multi-ancestry fine mapping for the two risk loci identified in both the TUD-EUR and TUD-AA metaGWAS. For each locus, we extracted SNPs with an absolute value of Z-score larger than 3.9 within a 1Mb region of the lead SNP. As suggested by PAINTOR, we created the AA and EUR LD matrices using the 1000 Genome phase 3 reference panel^77^. We calculated the probability of each SNP being the causal variant, assuming that each locus has two causal variants.

### Gene-based and pathway analyses

We conducted bioannotation and bioinformatic analyses to further characterize the loci identified by the TUD GWAS (**Supplementary Methods**). We used the default version (v1.3.6a) of the FUMA web-based platform^41^ to identify independent SNPs (*r*^2^<0.10) and to study their functional consequences. We also used MAGMA v1.08^41,42^ to perform competitive gene-set and pathway analyses. SNPs were mapped to 19,532 protein-coding genes from Ensembl (build 85). We applied a Bonferroni correction based on the total number of genes tested (*p*<2.63E−06). Gene sets were obtained from Msigdb v7.0 (“Curated gene sets”, “GO terms”). We also used Hi-C coupled MAGMA (H-MAGMA^43^) to assign non-coding (intergenic and intronic) SNPs to genes based on their chromatin interactions. Exonic and promoter SNPs were assigned to genes based on physical position. H-MAGMA uses four Hi-C datasets, which were derived from fetal brain, adult brain, iPSC-derived neurons, and iPSC-derived astrocytes (https://github.com/thewonlab/H-MAGMA). We applied a Bonferroni correction based on the total number of gene-tissue pairs tested (*p*<9.44E−07).

### S-MultiXcan/S-PrediXcan

We used S-MultiXcan v0.7.0 (an extension of S-PrediXcan v0.6.2^44^) to identify specific eQTL-linked genes associated with TUD. This approach uses genetic information to predict transcript abundance in 13 brain tissues, and tests whether the predicted transcripts correlate with TUD. S-PrediXcan uses pre-computed tissue weights from the Genotype-Tissue Expression (GTEx) v8 project database (https://www.gtexportal.org/) as the reference transcriptome dataset. For S-PrediXcan and S-MultiXcan analyses, we chose to use sparse (elastic net) prediction models, which are available at http://predictdb.hakyimlab.org/. We applied a conservative Bonferroni correction based on the total number of gene-tissue pairs tested (14,198 gene-tissue pairs tested; *p*<3.52E−06).

### PWAS/TWAS

To identify proteins whose genetically regulated expression is associated with TUD, we performed PWAS analyses by integrating TUD GWAS summary statistics and precomputed pQTLs from discovery (Banner)^98,99^ and validation (ROSMAP)^100,101^ datasets using the FUSION pipeline (http://gusevlab.org/projects/fusion/).^45^ Next, TWAS was performed using gene and splicing expression profiles measured in the adult DLPFC and gene expression profiles from the frontal cortex. Human brain transcriptome data, used as expression reference panels, were obtained from the CMC^100^ and GTEx frontal cortex v7.^45,95^ All tests were Bonferroni corrected for multiple testing (α = 0.05/N genes tested).

### Partitioning Heritability Enrichment

We used LDSC to partition TUD-EUR *h^2^_SNP_* and examined the enrichment based on several functional genomic annotation models.^92,93^ In the baseline model, we examined 75 overlapping functional annotations comprising genomic, epigenomic and regulatory features. We also analyzed ten overlapping cell-type groups derived from 220 cell-type-specific annotations in four histone marks: methylated histone H3 Lys4 (H3K4me1), trimethylated histone H3 Lys4 (H3K4me3), acetylated histone H3 Lys4 (H3K4ac) and H3K27ac. Enriched cell-type categories were analyzed based on annotations obtained from H3K4me1-imputed, gapped peak data generated by the Roadmap Epigenomics Mapping Consortium.^94^ We removed multi-allelic and major histocompatibility complex region variants, and only report categories enriched after Bonferroni correction.

### Tissue Enrichment Analysis

We used the LDSC package to conduct cell type specific heritability analysis (https://www.nature.com/articles/s41588-018-0081-4). In this analysis, we applied stratified LD score regression on the TUD-EUR meta-analysis summary statistics with sets of specifically expressed genes in various tissues from GTEx^95–97^ to identify TUD-relevant tissues. We applied a conservative Bonferroni correction based on the number of tissues simultaneously tested (205 tissues tested, *p*<2.44E-04). We also used MAGMA v1.08 gene-property analysis of expression data from GTEx (54 tissue types) and BrainSpan (29 brain samples at different age) in FUMA v1.3.6a^85^ to test the relationships between tissue specific gene expression profiles and TUD-gene associations.

### Cell type-specific expression of TUD risk genes

We performed cell-type specific analyses implemented in FUMA, using data from nine single-cell RNA sequencing data sets from human brain (data sets listed in the **Supplementary Material**). The method is described in detail in Watanabe et al.,^41^ and uses MAGMA gene-property analysis to test for association between cell specific gene expression and TUD-gene association. Conditional analyses for multiple testing are applied to correct for all tested cell types across datasets.

Of the overlapping findings across independent TWAS or PWAS datasets, colocalization analysis (in FUSION^45,102^) was used to determine whether SNPs mediate the association with TUD via effects on gene and protein expression. A posterior colocalization probability (PP) of 80% was used to indicate a shared causal signal.

### BrainXcan

We used the BrainXcan package (https://github.com/hakyimlab/brainxcan)^103^ to predict the association between the TUD phenotype and brain features. This approach uses genetically determined brain image-derived phenotypes (IDPs) to test brain region association with the TUD phenotype via linear regression. IDPs were constructed by training genetic predictors on original IDPs from MRI images via ridge regression.^103^ IDPs were retrieved from the BrainXcan database (https://zenodo.org/record/4895174). Only significant IDP associations with TUD that survived a Bonferroni correction are reported (93 IDPs tested; *p*<1.92E-04).

### Drug repurposing

Our signature matching technique used data from the Library of Integrated Network-based Cellular Signatures (**LINCs**) L1000 database. The LINCs L1000 database catalogues in vitro gene expression profiles (signatures) from thousands of compounds in over 80 human cell lines (level 5 data from phase I: GSE92742 and phase II: GSE70138). We selected compounds that were currently FDA approved or in clinical trials (via https://clue.io/repurposing#download-data; updated 3/24/20). Our analyses included signatures of 829 chemical compounds (590 FDA approved, 239 in clinical trials) in five neuronal cell-lines (NEU, NPC, MNEU.E, NPC.CAS9 and NPC.TAK), a total of 3,897 signatures.

We matched in vitro medication signatures with TUD signatures from brain tissue transcriptome-wide association analyses (conducted using S-PrediXcan). This consisted of Amygdala, Anterior Cingulate Cortex BA24, Caudate Basal Ganglia, Cerebellar Hemisphere, Cerebellum, Cortex, Frontal Cortex BA9, Hippocampus, Hypothalamus, Nucleus Accumbens Basal Ganglia, Putamen Basal Ganglia, Substantia Nigra, and Pituitary brain regions. As previously described,^36^ we computed weighted Pearson correlations between transcriptome-wide brain associations and in vitro L1000 compound signatures, weighting each gene by its proportion of heritability explained, using the *metafor* package (version 3.8-1) in R. We treated each L1000 compound as a fixed effect incorporating the effect size (rweighted) and sampling variability (se2r_weighted) from all signatures of a compound (e.g., across all time points, cell lines, doses). Brain region was included as a random effect to account for any tissue specific heterogeneity. Both the genes for the transcriptome wide association analysis input and the medications from our drug repurposing analyses were required to survive a Bonferroni correction for multiple testing (transcriptome-wide correction=0.05/14,199=3.52E-06; Perturbagen correction =0.05/3,897=1.28E-05).

We applied an additional drug repositioning method, DRUGSETS.^104^ Data were drawn from the Clue Repurposing Hub and the Drug Gene Interaction Database. Drug gene-sets were created for 1,201 drugs with genes whose protein products are targeted by or interact with that specific drug. Competitive gene-set analysis was performed using MAGMA v1.08^41,42^ while conditioning on a gene set of all drug target genes in the data (N=2,281) to test for significant associations between drug-gene sets and TUD. We applied a Bonferroni correction for the number of drug-gene sets tested (*p*<0.05/735=6.80E-05).

### Genetic correlation analyses

We estimated the within-ancestry *r_g_s* for TUD using LDSC^46^ and the cross-ancestry *r_g_s* for TUD across population groups using POPCORN.^46^ We used the ancestry-specific 1000 Genomes Project phase 3^78^ data as the LD references.

We used local LDSC^46^ to calculate genetic correlations (*r_g_*) between TUD and 113 other traits or diseases.^46^ Local traits were selected based on previously known phenotypic associations between TUD and other substance use disorder phenotypes and related traits (e.g., cannabis use disorder, various measures of impulsivity). We used the standard Benjamini–Hochberg false discovery rate correction (FDR 5%) to correct for multiple testing. We also calculated a Bonferroni correction for 113 comparisons (*p*<4.42E−04); however, this correction is overly conservative because many of the traits tested are highly correlated with one another. For AA individuals, we calculated *r_g_* between TUD and 11 published traits using in-sample LD scores derived from the MVP genotype data using cov-LDSC.^87^

### mtCOJO

We used mtCOJO^105^ to individually condition the TUD-EUR summary statistics on loci associated with other comorbid traits, including alcohol dependence, cannabis use disorder and opioid use disorder. This analysis allowed us to examine whether the genetic associations with TUD would be preserved when controlling for those covariate phenotypes. To test as many SNPs while preserving computational efficiency, we used a *p* value threshold of 5.00E-06, 5.00E-08, 5.00E-06, respectively, for alcohol dependence, cannabis use disorder, and opioid use disorder. We then computed genetic correlations using the TUD summary statistics adjusted for the covariates of interest.

### Unsupervised learning to determine TUD clustering

Previous studies have shown that consumption and misuse/dependence phenotypes have a distinct genetic architecture. To explore whether the TUD meta-analysis clustered more with consumption or misuse/dependence phenotypes, we used a data-driven unsupervised machine learning method known as agglomerative hierarchical clustering analysis (**HCA**).^106^ HCA forms clusters iteratively by creating groups and successively joining or splitting those groups based on a prespecified algorithm.^106^ Agglomerative nesting (AGNES) is a bottom-up process focused on individual traits to structure. Agglomerative clustering was chosen as this allowed us to compare different algorithms to maximize for the dissimilarity on each branch, with Ward’s minimum variance method performing best. All models were fit in R using the *cluster* package (version 2.1.4).^106^

The product of HCA is a dendrogram, formed with multiple brackets called “branches”. Phenotypes on the same branch are more similar to each other based on their pairwise genetic associations with each other and with all other phenotypes on that branch. Branches can form subbranches of more specific clustering. The genetic correlations of Former Smoker and Smoking Initiation were reversed to show the intuitive effects against the other traits in the dendrogram.

### Phenome-wide association studies (PheWAS)

#### Mayo Clinic Biobank

We performed a PheWAS in the Mayo Clinic Biobank (**MCB**).^107^ Phecodes were ascertained using EHR data from 57,001 patients from the Mayo Clinic Biobank. EHR data for the participants was extracted on September 23, 2022 and included any diagnoses on or before April 6, 2020, the date patient consent was checked. The Institutional Review Board of Mayo Clinic approved this study. Samples were sequenced at the Regeneron Genetics Center (**RGC**) using a custom design that additionally augments the exome capture with “backbone” regions intended to measure common tagging variation for purposes of GWAS. The backbone regions are targeted at lower depth and undergo substantial post-processing using proprietary algorithms that can boost genotyping quality based on shared information via linkage disequilibrium and population allele frequencies. The resulting GxS data was run through the Mayo Clinic Genotype QC pipeline. SNPs were excluded using filters for call rate (<95%), minor allele frequency (<0.5%), and Hardy-Weinberg Equilibrium (*p*<1.00E-06). Individuals were excluded for excessive missing genotypes (>5%), sex errors, or abnormal heterozygosity (<70% on multiple chromosomes). Cryptic relatedness analysis was performed in an iterative process using PLINK and PRIMUS to estimate IBD sharing. Highly related samples were removed from the sample if they had >100 closely related samples (PI_HAT>0.1875) or >25000 related samples (PI_HAT>0.08); the relatedness analysis was performed iteratively until no such samples remained. For each pair with an estimated 2^nd^ degree or higher relatedness, we removed the individual with shorter length of EHR. PGS were calculated using LDpred2^108^ l using the auto feature in the bigsnpr (v1.10.4) R package. To evaluate the unique contribution of polygenic scores for TUD in relation to other smoking behaviors, we calculated PGS for SmkInit, CPD^13^ and FTND^27^ and ran additional PheWAS of TUD covarying for SmikInit, CPD and FTND PGS.

#### Yale-Penn

We performed PheWAS in the Yale-Penn sample;^47^ which is a deeply phenotyped cohort using the Semi-Structured Assessment for Drug Dependence and Alcoholism, a detailed psychiatric instrument used to assess physical, psychosocial, and psychiatric manifestations of SUDs and comorbid psychiatric traits.^109,110^ This comprehensive interview includes more than 3,500 items representing lifetime diagnostic criteria for the DSM-IV,^111^ DSM-5^112^ SUDs and DSM-IV^111^ psychiatric disorder history. Genotyping and quality control for this cohort have been extensively described.^47,113^

PGSs were calculated using PRS-Continuous shrinkage software (PRS-CS).^114^ We used the default setting in PRS-CS to estimate the shrinkage parameters and fixed the random seed to 1 for reproducibility. To identify associations between the PGS for TUD and clinical phenotypes, we performed a PheWAS by fitting logistic regression models for binary phenotypes and linear regression models for continuous phenotypes. Analyses were conducted using the PheWAS v0.12 R package^115^ adjusting for sex, median age and the first ten PCs within each genetic ancestry. We performed sensitivity analyses by covarying for SmkInit, CPD^13^ and FTND^27^ PGS. Bonferroni correction was applied for each ancestral-specific analysis to account for multiple testing (*p*<7.25E-05).

#### Adolescent Brain Cognitive Development (ABCD)

We performed polygenic analyses in the ABCD sample.^116^ Again using PRS-CS,^117^ we fitted a fixed effects model in the ABCD European subsample (wave 3 for phenotypes, wave 3 for genotypes), controlling for first 10 PCs, age, sex, site, as fixed effect covariates and family ID as random effects covariates. We included 12 measures that showed significant *rg* in the adults datasets and were available in this cohort; these included 2 binary phenotypes (pain, “any pain last month”; and suicide attempt, “description”), and 10 continuous measures (from the CBCL child behavior checklist>^118^– “CBCL Externalizing”, “CBCL ADHD”, “CBCL Depression”, “CBCL ADHD”, “CBCL AnxDep”; “CBCL AnxDis”, “CBCL OCD”; cognitive ability via the NIH cognitive toolbox total score;^119^ BMI; weight; deprivation). Results were corrected for multiple testing (*p*<4.0E-03). Additional genotyping, QC and statistical details are described in the **Supplementary Material**.

### Mendelian Randomization

Two-sample Mendelian randomization^120,121^ was used to evaluate the potential causal association between TUD and genetically correlated traits using samples of European ancestry only (without UKBB). Of the 76 traits that showed significant genetic correlations (**Supplementary Table 31**), we removed 45 that were phenotypically similar (e.g., BMI and obesity). From each category, we selected those traits with higher *r_g_*. Therefore, we tested 31 traits for a causal relationship with TUD. We inferred causality bidirectionally using three methods: weighted median, inverse-variance weighted (**IVW**) and MR-Egger, followed by a pleiotropy test using the MR Egger intercept.^122,123^ Instrumental variants were those associated with the exposure after clumping (*r*^2^ = 0.01) and at *p*<1.0E-05. We considered causal effects as those for which at least two MR tests were significant after Bonferroni correction (*p* = 0.05/31 = 1.61E-03) and that showed no evidence of violation of the horizontal pleiotropy test (MR-Egger intercept *p*>0.05).

## Data Availability

The full summary statistics from the meta-analyses will be available through dbGaP upon publication.

## Code Availability

All software used to generate results has been previously published, and corresponding citations are provided in the Methods.

## Acknowledgements

MVJ, SBB, SRP and SSR were supported by funds from the California Tobacco-Related Disease Research Program (TRDRP; Grant Number T29KT0526 and T32IR5226). SBB and were also supported by P50DA037844. BKP, JM and SSR were supported by NIH/NIDA DP1DA054394. ASH was supported by NIAAA AA030083. TTM was supported by NHGRI T32HG010464. ECJ was supported by K01DA051759. JG was supported by VA Merit Award CX001849-01 and 5R01DA054869. DBH was supported by R01 DA042090. LKD was supported by R01 MH113362. HRK was supported by the Veterans Integrated Service Network 4 Mental Illness Research, Education and Clinical Center. RLK was supported by NIAAA K01 AA028292. The content is solely the responsibility of the authors and does not necessarily represent the official views of the National Institutes of Health.

## CTSA (SD, Vanderbilt Resources)

The project described was supported by the National Center for Research Resources, Grant UL1 RR024975-01, and is now at the National Center for Advancing Translational Sciences, Grant 2 UL1 TR000445-06.

## BioVU

The dataset(s) used for the analyses described were obtained from Vanderbilt University Medical Center’s BioVU which is supported by numerous sources: institutional funding, private agencies, and federal grants. These include the NIH funded Shared Instrumentation Grant S10RR025141; and CTSA grants UL1TR002243, UL1TR000445, and UL1RR024975. Genomic data are also supported by investigator-led projects that include U01HG004798, R01NS032830, RC2GM092618, P50GM115305, U01HG006378, U19HL065962, R01HD074711; and additional funding sources listed at https://victr.vumc.org/biovu-funding/.

This research is based on data from the Million Veteran Program, Office of Research and Development, Veterans Health Administration, and was supported by funding from the Department of Veterans Affairs Office of Research and Development, Million Veteran Program Grant #I01 BX004820. This publication does not represent the views of the Department of Veterans Affairs or the United States Government.

We acknowledge the Penn Medicine BioBank (PMBB) and the Mayo Clinic Biobank for providing data and thank the patient-participants of Penn Medicine and Mayo Clinic who consented to participate in this research program. We would also like to thank the Penn Medicine BioBank team and Regeneron Genetics Center for providing genetic variant data for analysis. The PMBB is approved under IRB protocol# 813913 and supported by Perelman School of Medicine at University of Pennsylvania, a gift from the Smilow family, and the National Center for Advancing Translational Sciences of the National Institutes of Health under CTSA award number UL1TR001878.

Data used in the preparation of this article were obtained from the Adolescent Brain Cognitive Development (ABCD) Study (https://abcdstudy.org), held in the NIMH Data Archive (NDA). This is a multisite, longitudinal study designed to recruit more than 10,000 children age 9-10 and follow them over 10 years into early adulthood. The ABCD Study is supported by the National Institutes of Health and additional federal partners under award numbers U01DA041022, U01DA041028, U01DA041048, U01DA041089, U01DA041106, U01DA041117, U01DA041120, U01DA041134, U01DA041148, U01DA041156, U01DA041174, U24DA041123, U24DA041147, U01DA041093, and U01DA041025. A full list of supporters is available at https://abcdstudy.org/federal-partners.html. A listing of participating sites and a complete listing of the study investigators can be found at https://abcdstudy.org/Consortium_Members.pdf. ABCD consortium investigators designed and implemented the study and/or provided data but did not necessarily participate in analysis or writing of this report. This manuscript reflects the views of the authors and may not reflect the opinions or views of the NIH or ABCD consortium investigators.

We would also like to thank The Externalizing Consortium for sharing the GWAS summary statistics of externalizing. The Externalizing Consortium: Principal Investigators: Danielle M. Dick, Philipp Koellinger, K. Paige Harden, Abraham A. Palmer. Lead Analysts: Richard Karlsson Linnér, Travis T. Mallard, Peter B. Barr, Sandra Sanchez-Roige. Significant Contributors: Irwin D. Waldman. The Externalizing Consortium has been supported by the National Institute on Alcohol Abuse and Alcoholism (R01AA015416 -administrative supplement), and the National Institute on Drug Abuse (R01DA050721). Additional funding for investigator effort has been provided by K02AA018755, U10AA008401, P50AA022537, as well as a European Research Council Consolidator Grant (647648 EdGe to Koellinger). The content is solely the responsibility of the authors and does not necessarily represent the official views of the above funding bodies. The Externalizing Consortium would like to thank the following groups for making the research possible: 23andMe, Add Health, Vanderbilt University Medical Center’s BioVU, Collaborative Study on the Genetics of Alcoholism (COGA), the Psychiatric Genomics Consortium’s Substance Use Disorders working group, UK10K Consortium, UK Biobank, and Philadelphia Neurodevelopmental Cohort.

## Contributions

The corresponding author (S.S-R) conceived the idea for the paper and wrote and edited the manuscript. Other contributing authors contributed analyses (S.T., M.V.J., B.P., H.L., T.T.L., S.B.B., L.V-R, H.X., A.H., J.J.M., V.P., G.Y., B.S.L., B.C., R.L.K), or data (E.C.J., MVP, G.D.J., A.B., R.P., R.B., A.A., J.B., J.W.S., L.K.D., A.C.J., R.L.K.). All contributing authors wrote and edited the paper. We thank Bryan Quach and Jesse Marks for their help in supplying portions of the data needed to create Supplementary Figure 1.

## Ethics declarations

Dr. Palmer is on the scientific advisory board of Vivid Genomics for which he receives stock options. Dr. Smoller is a member of the Scientific Advisory Board of Sensorium Therapeutics (with equity) and has received grant support from Biogen, Inc. He is PI of a collaborative study of the genetics of depression and bipolar disorder sponsored by 23andMe for which 23andMe provides analysis time as in-kind support but no payments. Dr. Kranzler is a member of advisory boards for Clearmind Medicine, Dicerna Pharmaceuticals, Sophrosyne Pharmaceuticals, and Enthion Pharmaceuticals; a consultant to Sobrera Pharmaceuticals; the recipient of research funding and medication supplies for an investigator-initiated study from Alkermes; a member of the American Society of Clinical Psychopharmacology’s Alcohol Clinical Trials Initiative, which was supported in the last three years by Alkermes, Dicerna, Ethypharm, Lundbeck, Mitsubishi, Otsuka, and Pear Therapeutics; and with Dr. Gelernter, a holder of U.S. patent 10,900,082 titled: “Genotype-guided dosing of opioid agonists,” issued 26 January 2021. The other authors declare no competing interests.

## References

1. Centers for Disease Control and Prevention (CDC). Health Effects of Cigarette Smoking. https://www.cdc.gov/tobacco/data_statistics/fact_sheets/health_effects/effects_cig_smoking/index.htm (2021).

2. Oliver, J. A. & Foulds, J. Association Between Cigarette Smoking Frequency and Tobacco Use Disorder in U.S. Adults. Am. J. Prev. Med. 60, 726–728 (2021).

3. WHO. The top 10 causes of death. World Health Organization https://www.who.int/news-room/fact-sheets/detail/the-top-10-causes-of-death.

4. Benowitz, N. L. & Liakoni, E. Tobacco use disorder and cardiovascular health. Addiction 117, 1128–1138 (2022).

5. Kalman, D., Morissette, S. B. & George, T. P. Co-Morbidity of Smoking in Patients with Psychiatric and Substance Use Disorders. Am. J. Addict. Am. Acad. Psychiatr. Alcohol. Addict. 14, 106–123 (2005).

6. Tobacco use disorder and the lungs - McRobbie - 2021 - Addiction - Wiley Online Library. https://onlinelibrary.wiley.com/doi/10.1111/add.15309.

7. Ziedonis, D., Das, S. & Larkin, C. Tobacco use disorder and treatment: new challenges and opportunities. Dialogues Clin. Neurosci. 19, 271–280 (2017).

8. Kendler, K. S., Schmitt, E., Aggen, S. H. & Prescott, C. A. Genetic and Environmental Influences on Alcohol, Caffeine, Cannabis, and Nicotine Use From Early Adolescence to Middle Adulthood. Arch. Gen. Psychiatry 65, 674–682 (2008).

9. Do, E. K. et al. Genetic and Environmental Influences on Smoking Behavior across Adolescence and Young Adulthood in the Virginia Twin Study of Adolescent Behavioral Development and the Transitions to Substance Abuse Follow-Up. Twin Res. Hum. Genet. Off. J. Int. Soc. Twin Stud. 18, 43–51 (2015).

10. Agrawal, A., Budney, A. J. & Lynskey, M. T. The Co-occurring Use and Misuse of Cannabis and Tobacco: A Review. Addict. Abingdon Engl. 107, 1221–1233 (2012).

11. Agrawal, A. et al. The genetics of addiction—a translational perspective. Transl. Psychiatry 2, e140–e140 (2012).

12. Sullivan, P. F. & Kendler, K. S. The genetic epidemiology of smoking. Nicotine Tob. Res. 1, S51–S57 (1999).

13. Saunders, G. R. B. et al. Genetic diversity fuels gene discovery for tobacco and alcohol use. Nature 612, 720–724 (2022).

14. Larsson, S. C. & Burgess, S. Appraising the causal role of smoking in multiple diseases: A systematic review and meta-analysis of Mendelian randomization studies. eBioMedicine 82, (2022).

15. Yuan, S., Michaëlsson, K., Wan, Z. & Larsson, S. C. Associations of Smoking and Alcohol and Coffee Intake with Fracture and Bone Mineral Density: A Mendelian Randomization Study. Calcif. Tissue Int. 105, 582–588 (2019).

16. Mahedy, L. et al. Testing the association between tobacco and cannabis use and cognitive functioning: Findings from an observational and Mendelian randomization study. Drug Alcohol Depend. 221, 108591 (2021).

17. Zhou, H. et al. Association of *OPRM1* Functional Coding Variant With Opioid Use Disorder: A Genome-Wide Association Study. JAMA Psychiatry 77, 1072 (2020).

18. Wootton, R. E. et al. Evidence for causal effects of lifetime smoking on risk for depression and schizophrenia: a Mendelian randomisation study. Psychol. Med. 50, 2435–2443 (2020).

19. Harrison, R., Munafò, M. R., Davey Smith, G. & Wootton, R. E. Examining the effect of smoking on suicidal ideation and attempts: triangulation of epidemiological approaches. Br. J. Psychiatry 217, 701–707.

20. Xu, K. et al. Genome-wide association study of smoking trajectory and meta-analysis of smoking status in 842,000 individuals. Nat. Commun. 11, 5302 (2020).

21. Sanchez-Roige, S. et al. Genome-wide association study of alcohol use disorder identification test (AUDIT) scores in 20 328 research participants of European ancestry: GWAS of AUDIT. Addict. Biol. 24, 121–131 (2019).

22. Kranzler, H. R. et al. Genome-wide association study of alcohol consumption and use disorder in 274,424 individuals from multiple populations. Nat. Commun. 10, 1499 (2019).

23. Mallard, T. T. & Sanchez-Roige, S. Dimensional Phenotypes in Psychiatric Genetics: Lessons from Genome-Wide Association Studies of Alcohol Use Phenotypes. Complex Psychiatry 7, 45–48 (2021).

24. Mallard, T. T. et al. Item-Level Genome-Wide Association Study of the Alcohol Use Disorders Identification Test in Three Population-Based Cohorts. Am. J. Psychiatry appi.ajp.2020.2 (2021) doi:10.1176/appi.ajp.2020.20091390.

25. Sanchez-Roige, S. & Palmer, A. A. Emerging phenotyping strategies will advance our understanding of psychiatric genetics. Nat. Neurosci. 23, 475–480 (2020).

26. Johnson, E. C. et al. A large-scale genome-wide association study meta-analysis of cannabis use disorder. Lancet Psychiatry 7, 1032–1045 (2020).

27. Quach, B. C. et al. Expanding the genetic architecture of nicotine dependence and its shared genetics with multiple traits. Nat. Commun. 11, 5562 (2020).

28. Hancock, D. B., Markunas, C. A., Bierut, L. J. & Johnson, E. O. Human Genetics of Addiction: New Insights and Future Directions. Curr. Psychiatry Rep. 20, 8 (2018).

29. Sanchez-Roige, S., Cox, N. J., Johnson, E. O., Hancock, D. B. & Davis, L. K. Alcohol and cigarette smoking consumption as genetic proxies for alcohol misuse and nicotine dependence. Drug Alcohol Depend. 221, 108612 (2021).

30. DeBoever, C. et al. Assessing Digital Phenotyping to Enhance Genetic Studies of Human Diseases. Am. J. Hum. Genet. 106, 611–622 (2020).

31. Sanchez-Roige, S. & Palmer, A. A. Electronic Health Records Are the Next Frontier for the Genetics of Substance Use Disorders. Trends Genet. 35, 317–318 (2019).

32. Zheutlin, A. B. et al. Penetrance and Pleiotropy of Polygenic Risk Scores for Schizophrenia in 106,160 Patients Across Four Health Care Systems. Am. J. Psychiatry 176, 846–855 (2019).

33. Verma, A. et al. The Penn Medicine BioBank: Towards a Genomics-Enabled Learning Healthcare System to Accelerate Precision Medicine in a Diverse Population. J. Pers. Med. 12, 1974 (2022).

34. Roughley, S., Marcus, A. & Killcross, S. Dopamine D1 and D2 Receptors Are Important for Learning About Neutral-Valence Relationships in Sensory Preconditioning. Front. Behav. Neurosci. 15, (2021).

35. Gelernter, J. et al. Haplotype spanning TTC12 and ANKK1, flanked by the DRD2 and NCAM1 loci, is strongly associated to nicotine dependence in two distinct American populations. Hum. Mol. Genet. 15, 3498–3507 (2006).

36. Hatoum, A. S., et al. Multivariate genome-wide association meta-analysis of over 1 million subjects identifies loci underlying multiple substance use disorders. http://medrxiv.org/lookup/doi/10.1101/2022.01.06.22268753 (2022) doi:10.1101/2022.01.06.22268753.

37. Liu, M. et al. Association studies of up to 1.2 million individuals yield new insights into the genetic etiology of tobacco and alcohol use. Nat. Genet. 51, 237–244 (2019).

38. Sanchez-Roige, S. et al. Genome-wide association study of problematic opioid prescription use in 132,113 23andMe research participants of European ancestry. Mol. Psychiatry 26, 6209–6217 (2021).

39. Karlsson Linnér, R., et al. Multivariate analysis of 1.5 million people identifies genetic associations with traits related to self-regulation and addiction. Nat. Neurosci. (2021) doi:10.1038/s41593-021-00908-3.

40. Xiao, M.-F. et al. Neural Cell Adhesion Molecule Modulates Dopaminergic Signaling and Behavior by Regulating Dopamine D2 Receptor Internalization. J. Neurosci. 29, 14752– 14763 (2009).

41. Watanabe, K., Umićević Mirkov, M., de Leeuw, C. A., van den Heuvel, M. P. & Posthuma, D. Genetic mapping of cell type specificity for complex traits. Nat. Commun. 10, 3222 (2019).

42. Leeuw, C. A. de, Mooij, J. M., Heskes, T. & Posthuma, D. MAGMA: Generalized Gene-Set Analysis of GWAS Data. PLOS Comput. Biol. 11, e1004219 (2015).

43. Sey, N. Y. A. et al. A computational tool (H-MAGMA) for improved prediction of brain-disorder risk genes by incorporating brain chromatin interaction profiles. Nat. Neurosci. 23, 583–593 (2020).

44. Barbeira, A. N. et al. Exploring the phenotypic consequences of tissue specific gene expression variation inferred from GWAS summary statistics. Nat. Commun. 9, 1825 (2018).

45. Gusev, A. et al. Integrative approaches for large-scale transcriptome-wide association studies. Nat. Genet. 48, 245–252 (2016).

46. Bulik-Sullivan, B. K. et al. LD Score regression distinguishes confounding from polygenicity in genome-wide association studies. Nat. Genet. 47, 291–295 (2015).

47. Kember, R. L. et al. Phenome-wide Association Analysis of Substance Use Disorders in a Deeply Phenotyped Sample. Biol. Psychiatry (2022) doi:10.1016/j.biopsych.2022.08.010.

48. Sanchez-Roige, S., Palmer, A. A. & Clarke, T.-K. Recent Efforts to Dissect the Genetic Basis of Alcohol Use and Abuse. Biol. Psychiatry 87, 609–618 (2020).

49. McLellan, A. T., Koob, G. F. & Volkow, N. D. Preaddiction—A Missing Concept for Treating Substance Use Disorders. JAMA Psychiatry 79, 749–751 (2022).

50. Brazel, D. M. et al. Exome Chip Meta-analysis Fine Maps Causal Variants and Elucidates the Genetic Architecture of Rare Coding Variants in Smoking and Alcohol Use. Biol. Psychiatry 85, 946–955 (2019).

51. Miranda, M., Morici, J. F., Zanoni, M. B. & Bekinschtein, P. Brain-Derived Neurotrophic Factor: A Key Molecule for Memory in the Healthy and the Pathological Brain. Front. Cell. Neurosci. 13, (2019).

52. Barker, J. M., Taylor, J. R., De Vries, T. J. & Peters, J. Brain-derived neurotrophic factor and addiction: Pathological versus therapeutic effects on drug seeking. Brain Res. 1628, 68–81 (2015).

53. Duong, C. et al. Glutathione peroxidase-1 protects against cigarette smoke-induced lung inflammation in mice. Am. J. Physiol.-Lung Cell. Mol. Physiol. 299, L425–L433 (2010).

54. Scieszka, D. et al. Subchronic Electronic Cigarette Exposures Have Overlapping Protein Biomarkers with Chronic Obstructive Pulmonary Disease and Idiopathic Pulmonary Fibrosis. Am. J. Respir. Cell Mol. Biol. 67, 503–506 (2022).

55. Aberg, K. A. et al. A Comprehensive Family-Based Replication Study of Schizophrenia Genes. JAMA Psychiatry 70, 573 (2013).

56. Erzurumluoglu, A. M. et al. Meta-analysis of up to 622,409 individuals identifies 40 novel smoking behaviour associated genetic loci. Mol. Psychiatry 25, 2392–2409 (2020).

57. Toikumo, S., Xu, H., Gelernter, J., Kember, R. L. & Kranzler, H. R. Integrating human brain proteomic data with genome-wide association study findings identifies novel brain proteins in substance use traits. Neuropsychopharmacology 47, 2292–2299 (2022).

58. Kember, R. L. et al. Cross-ancestry meta-analysis of opioid use disorder uncovers novel loci with predominant effects in brain regions associated with addiction. Nat. Neurosci. 25, 1279–1287 (2022).

59. Koob, G. F. & Volkow, N. D. Neurobiology of addiction: a neurocircuitry analysis. Lancet Psychiatry 3, 760–773 (2016).

60. King, D. P. et al. Smoking Cessation Pharmacogenetics: Analysis of Varenicline and Bupropion in Placebo-Controlled Clinical Trials. Neuropsychopharmacology 37, 641–650 (2012).

61. King, A. C. et al. Effects of Naltrexone on Smoking Cessation Outcomes and Weight Gain in Nicotine-Dependent Men and Women. J. Clin. Psychopharmacol. 32, 630–636 (2012).

62. Carpenter, M. J. et al. Clinical Strategies to Enhance the Efficacy of Nicotine Replacement Therapy for Smoking Cessation: A Review of the Literature. Drugs 73, 407–426 (2013).

63. So, H.-C. et al. Analysis of genome-wide association data highlights candidates for drug repositioning in psychiatry. Nat. Neurosci. 20, 1342–1349 (2017).

64. Sey, N. Y. A. et al. Chromatin architecture in addiction circuitry identifies risk genes and potential biological mechanisms underlying cigarette smoking and alcohol use traits. Mol. Psychiatry 27, 3085–3094 (2022).

65. Chen, F. et al. Multi-ancestry transcriptome-wide association analyses yield insights into tobacco use biology and drug repurposing. Nat. Genet. 55, 291–300 (2023).

66. Jamali, Q. Galantamine as a Treatment Option for Nicotine Addiction. J. Smok. Cessat. 2021, 9975811 (2021).

67. McGinnis, K. A., et al. Using the biomarker cotinine and survey self-report to validate smoking data from United States Veterans Health Administration electronic health records. JAMIA Open 5, ooac040 (2022).

68. Border, R. et al. Cross-trait assortative mating is widespread and inflates genetic correlation estimates. Science 378, 754–761 (2022).

69. Jang, S.-K. et al. Rare genetic variants explain missing heritability in smoking. *Nat*. Hum. Behav. 6, 1577–1586 (2022).

70. Malhotra, D. & Sebat, J. CNVs: harbingers of a rare variant revolution in psychiatric genetics. Cell 148, 1223–1241 (2012).

71. Hiscock, R., Bauld, L., Amos, A., Fidler, J. A. & Munafò, M. Socioeconomic status and smoking: a review. Ann. N. Y. Acad. Sci. 1248, 107–123 (2012).

72. Pasman, J. A. et al. Genetic Risk for Smoking: Disentangling Interplay Between Genes and Socioeconomic Status. Behav. Genet. 52, 92–107 (2022).

73. Treur, J. L. et al. Testing Familial Transmission of Smoking With Two Different Research Designs. Nicotine Tob. Res. Off. J. Soc. Res. Nicotine Tob. 20, 836–842 (2018).

74. Meyers, J. L. et al. Interaction between polygenic risk for cigarette use and environmental exposures in the Detroit Neighborhood Health Study. Transl. Psychiatry 3, e290 (2013).

75. Pasman, J. A., Verweij, K. J. H. & Vink, J. M. Systematic Review of Polygenic Gene-Environment Interaction in Tobacco, Alcohol, and Cannabis Use. Behav. Genet. 49, 349– 365 (2019).

76. Sanchez-Roige, S., Kember, R. L. & Agrawal, A. Substance use and common contributors to morbidity: A genetics perspective. EBioMedicine 83, 104212 (2022).

77. The 1000 Genomes Project Consortium et al. A global reference for human genetic variation. Nature 526, 68–74 (2015).

78. Fang, H. et al. Harmonizing Genetic Ancestry and Self-identified Race/Ethnicity in Genome-wide Association Studies. Am. J. Hum. Genet. 105, 763–772 (2019).

79. Zhou, W. et al. Efficiently controlling for case-control imbalance and sample relatedness in large-scale genetic association studies. Nat. Genet. 50, 1335–1341 (2018).

80. Chang, C. C. et al. Second-generation PLINK: rising to the challenge of larger and richer datasets. GigaScience 4, 7 (2015).

81. Dennis, J. K. et al. Clinical laboratory test-wide association scan of polygenic scores identifies biomarkers of complex disease. Genome Med. 13, 6 (2021).

82. Purcell, S. et al. PLINK: A Tool Set for Whole-Genome Association and Population-Based Linkage Analyses. Am. J. Hum. Genet. 81, 559–575 (2007).

83. Das, S. et al. Next-generation genotype imputation service and methods. Nat. Genet. 48, 1284–1287 (2016).

84. Price, A. L. et al. Principal components analysis corrects for stratification in genome-wide association studies. Nat. Genet. 38, 904–909 (2006).

85. Delaneau, O., Zagury, J.-F., Robinson, M. R., Marchini, J. L. & Dermitzakis, E. T. Accurate, scalable and integrative haplotype estimation. Nat. Commun. 10, 5436 (2019).

86. Altshuler, D. M., Gibbs, R. A., Peltonen, L., Dermitzakis, E. T. & Schaffner, S. F. Integrating common and rare genetic variation in diverse human populations. Nature 467, 52–58 (2010).

87. Luo, Y. et al. Estimating heritability and its enrichment in tissue-specific gene sets in admixed populations. Human Molecular Genetics 30, 1521–1534 (2021).

88. Willer, C. J., Li, Y. & Abecasis, G. R. METAL: fast and efficient meta-analysis of genomewide association scans. Bioinformatics 26, 2190–2191 (2010).

89. Yang, J., Lee, S. H., Goddard, M. E. & Visscher, P. M. GCTA: A Tool for Genome-wide Complex Trait Analysis. Am. J. Hum. Genet. 88, 76–82 (2011).

90. Benner, C. et al. FINEMAP: efficient variable selection using summary data from genome-wide association studies. Bioinformatics 32, 1493–1501 (2016).

91. Kichaev, G. & Pasaniuc, B. Leveraging Functional-Annotation Data in Trans-ethnic Fine-Mapping Studies. Am. J. Hum. Genet. 97, 260–271 (2015).

92. Finucane, H. K. et al. Partitioning heritability by functional annotation using genome-wide association summary statistics. Nat. Genet. 47, 1228–1235 (2015).

93. Finucane, H. K. et al. Heritability enrichment of specifically expressed genes identifies disease-relevant tissues and cell types. Nat. Genet. 50, 621–629 (2018).

94. Bernstein, B. E. et al. The NIH Roadmap Epigenomics Mapping Consortium. Nat. Biotechnol. 28, 1045–1048 (2010).

95. The GTEx Consortium atlas of genetic regulatory effects across human tissues. https://www.science.org/doi/10.1126/science.aaz1776 doi:10.1126/science.aaz1776.

96. Fehrmann, R. S. N. et al. Gene expression analysis identifies global gene dosage sensitivity in cancer. Nat. Genet. 47, 115–125 (2015).

97. Pers, T. H. et al. Biological interpretation of genome-wide association studies using predicted gene functions. Nat. Commun. 6, 5890 (2015).

98. Beach, T. G. et al. Arizona Study of Aging and Neurodegenerative Disorders and Brain and Body Donation Program. Neuropathol. Off. J. Jpn. Soc. Neuropathol. 35, 354–389 (2015).

99. Wingo, T. S. et al. Brain proteome-wide association study implicates novel proteins in depression pathogenesis. Nat. Neurosci. 24, 810–817 (2021).

100. Wingo, A. P. et al. Integrating human brain proteomes with genome-wide association data implicates new proteins in Alzheimer’s disease pathogenesis. Nat. Genet. 53, 143–146 (2021).

101. Bennett, D. A. et al. Religious Orders Study and Rush Memory and Aging Project. J. Alzheimers Dis. JAD 64, S161–S189 (2018).

102. Giambartolomei, C. et al. Bayesian Test for Colocalisation between Pairs of Genetic Association Studies Using Summary Statistics. PLoS Genet. 10, e1004383 (2014).

103. Liang, Y. et al. BrainXcan identifies brain features associated with behavioral and psychiatric traits using large scale genetic and imaging data. 2021.06.01.21258159 Preprint at 10.1101/2021.06.01.21258159 (2022).

104. Bell, N., Uffelmann, E., Walree, E. van, Leeuw, C. de & Posthuma, D. Using genome-wide association results to identify drug repurposing candidates. 2022.09.06.22279660 Preprint at 10.1101/2022.09.06.22279660 (2022).

105. Gu, Z., Gu, L., Eils, R., Schlesner, M. & Brors, B. circlize implements and enhances circular visualization in R. Bioinformatics 30, 2811–2812 (2014).

106. Maechler,M., Rousseeuw, P., Struyf, A., Hubert, M., Hornik, K. Cluster: Cluster Analysis Basics and Extentions. (2013).

107. Bielinski, S. J. et al. Mayo Genome Consortia: A Genotype-Phenotype Resource for Genome-Wide Association Studies With an Application to the Analysis of Circulating Bilirubin Levels. Mayo Clin. Proc. 86, 606–614 (2011).

108. Privé, F., Arbel, J. & Vilhjálmsson, B. J. LDpred2: better, faster, stronger. Bioinformatics 36, 5424–5431 (2020).

109. Pierucci-Lagha, A. et al. Diagnostic reliability of the Semi-structured Assessment for Drug Dependence and Alcoholism (SSADDA). Drug Alcohol Depend. 80, 303–312 (2005).

110. Pierucci-Lagha, A. et al. Reliability of DSM-IV Diagnostic Criteria Using the Semi-Structured Assessment for Drug Dependence and Alcoholism (SSADDA). Drug Alcohol Depend. 91, 85–90 (2007).

111. American Psychiatric Association. Diagnostic and Statistical Manual of Mental Disorders (DSM-IV). (American Psychiatric Association, 1994).

112. American Psychiatric Association. Diagnostic and Statistical Manual of Mental Disorders (DSM-5). (American Psychiatric Association, 2013).

113. Gelernter, J. et al. Genome-wide association study of alcohol dependence:significant findings in African- and European-Americans including novel risk loci. Mol. Psychiatry 19, 41–49 (2014).

114. Ge, T., Chen, C.-Y., Ni, Y., Feng, Y.-C. A. & Smoller, J. W. Polygenic prediction via Bayesian regression and continuous shrinkage priors. Nat. Commun. 10, 1776 (2019).

115. Denny, J. C., Bastarache, L. & Roden, D. M. Phenome-Wide Association Studies as a Tool to Advance Precision Medicine. Annu. Rev. Genomics Hum. Genet. 17, 353–373 (2016).

116. Lam, M. et al. RICOPILI: Rapid Imputation for COnsortias PIpeLIne. Bioinformatics 36, 930–933 (2020).

117. Ruan, Y. et al. Improving Polygenic Prediction in Ancestrally Diverse Populations. medRxiv 21 (2021) 10.1101/2020.12.27.20248738.

118. Rescorla, L. et al. Behavioral/Emotional Problems of Preschoolers Caregiver/Teacher Reports From 15 Societies. J. Emot. Behav. Disord. 20, 68–81 (2012).

119. Akshoomoff, N. et al. NIH Toolbox Cognitive Function Battery (CFB): Composite Scores of Crystallized, Fluid, and Overall Cognition. Monogr. Soc. Res. Child Dev. 78, 119–132 (2013).

120. Yavorska, O. O. & Burgess, S. MendelianRandomization: an R package for performing Mendelian randomization analyses using summarized data. Int. J. Epidemiol. 46, 1734– 1739 (2017).

121. Lawlor, D. A., Harbord, R. M., Sterne, J. A. C., Timpson, N. & Davey Smith, G. Mendelian randomization: Using genes as instruments for making causal inferences in epidemiology. Stat. Med. 27, 1133–1163 (2008).

122. Burgess, S., Butterworth, A. & Thompson, S. G. Mendelian Randomization Analysis With Multiple Genetic Variants Using Summarized Data. Genet. Epidemiol. 37, 658–665 (2013).

123. Bowden, J., Davey Smith, G., Haycock, P. C. & Burgess, S. Consistent Estimation in Mendelian Randomization with Some Invalid Instruments Using a Weighted Median Estimator. Genet. Epidemiol. 40, 304–314 (2016).

