## Supplementary Material for "Multi-ancestry meta-analysis of tobacco use disorder prioritizes novel candidate risk genes and reveals associations with numerous health outcomes"

^1^Mental Illness Research, Education and Clinical Center, Crescenz VAMC, Philadelphia, PA, USA; ^2^Department of Psychiatry, University of Pennsylvania Perelman School of Medicine, Philadelphia, PA, USA; ^3^Department of Psychiatry, University of California San Diego, San Diego, CA, USA; ^4^Psychiatric and Neurodevelopmental Genetics Unit, Center for Genomic Medicine, Massachusetts General Hospital, Boston, MA, USA; ^5^Department of Psychiatry, Harvard Medical School, Boston, MA, USA; ^6^Psychiatric Genetics Unit, Group of Psychiatry, Mental Health and Addiction, Vall d’Hebron Research Institute (VHIR), Universitat Autònoma de Barcelona, Barcelona, Spain; ^7^Department of Psychiatry, Washington University School of Medicine, Saint Louis, Missouri, USA; ^8^Department of Quantitative Health Sciences, Mayo Clinic, Rochester, MN, USA; ^9^Program in Biomedical Sciences, University of California San Diego, La Jolla, CA, USA; ^10^Vanderbilt Genetics Institute, Vanderbilt University Medical Center, Nashville, TN, USA; ^11^Department of Psychology, University of Ibadan, Nigeria; ^12^Institute for Genomic Medicine, University of California San Diego, La Jolla, CA, USA; ^13^Department of Psychiatry, Yale University School of Medicine, New Haven, CT, USA; ^14^Veterans Affairs Connecticut Healthcare System, West Haven, CT, USA; ^15^Department of Psychiatry & Psychology, Mayo Clinic, Rochester, MN, USA; ^16^Behavioral and Urban Health Program, Behavioral Health and Criminal Justice Division, RTI International, Research Triangle Park, NC, USA; ^17^Department of Medicine, Division of Genetic Medicine, Vanderbilt University, Nashville, TN, USA; ^18^Department of Biomedical Informatics, Vanderbilt University Medical Center, Nashville, TN, USA; ^19^Department of Psychiatry and Behavioral Sciences, Vanderbilt University Medical Center, Nashville, TN, USA; ^20^Yale University School of Public Health, New Haven, CT, USA; ^21^Veterans Affairs Connecticut Healthcare System, West Haven, CT, USA; ^22^Yale University School of Medicine, New Haven, CT, USA.

#### **Contents**

#### 1. Additional authors and contributions………………………………………………………………2-7

2. Materials and Methods

2.1 Bioannotation and bioinformatic secondary analyses………………………………...8-9

2.2 Phenome-wide association analyses……………………………………………….....9-10

3. Supplementary Figures

3.1 Cross-site genetic correlation estimates…………………………………………......…11

3.2 Loci which overlap between the current study and previous smoking GWAS…..….12

3.3 Effect-effect plot of TUD-multi lead SNPs vs GSCAN traits…………………………..13

3.4 Heterogeneity plot of TUD-multi GWAS results………………………………………...14

3.5 Effect-effect plot of TUD-EUR lead SNPs across ancestries....……………...……….15

3.6 Heterogeneity Plot of TUD-EUR GWAS results …………………..…………………...16

3.7 Genes identified via TWAS and PWAS analyses …………………..………………….19

3.8 Partitioned heritability in LDSC ……………………………………...…………………...18

3.9 Hierarchical Clustering Analysis ……………………..……...…………………………...19

4. References……………………………………………………………………………………….……20

#### **Additional authors and contributions**

2

**Penn Medicine BioBank Banner Author List and Contribution Statements**

**PMBB Leadership Team**

Daniel J. Rader, M.D., Marylyn D. Ritchie, Ph.D., Michael D. Feldman M.D.

Contribution: All authors contributed to securing funding, study design and oversight. All authors reviewed the final version of the manuscript.

**Patient Recruitment and Regulatory Oversight**

JoEllen Weaver, Nawar Naseer, Ph.D., M.P.H., Afiya Poindexter, Ashlei Brock, Khadijah Hu-Sain, Yi-An Ko

Contributions: JW manages patient recruitment and regulatory oversight of study. NN manages participant engagement, assists with regulatory oversight, and researcher access. AP, AB, KH, YK perform recruitment and enrollment of study participants.

**Lab Operations**

JoEllen Weaver, Meghan Livingstone, Fred Vadivieso, Ashley Kloter, Stephanie DerOhannessian, Teo Tran, Linda Morrel, Ned Haubein, Joseph Dunn

Contribution: JW, ML, FV, SD conduct oversight of lab operations. ML, FV, AK, SD, TT, LM perform sample processing. NH, JD are responsible for sample tracking and the laboratory information management system.

**Clinical Informatics**

Anurag Verma, Ph.D., Colleen Morse, M.S., Marjorie Risman, M.S., Renae Judy, B.S.

Contribution: All authors contributed to the development and validation of clinical phenotypes used to identify study subjects and (when applicable) controls.

**Genome Informatics**

Anurag Verma Ph.D., Shefali S. Verma, Ph.D., Yuki Bradford, M.S., Scott Dudek, M.S., Theodore Drivas, M.D., PH.D.

Contribution: A.V., S.S.V. are responsible for the analysis, design, and infrastructure needed to quality control genotype and exome data. Y.B. performs the analysis. T.D. and A.V. provides variant and gene annotations and their functional interpretation of variants.

**VA Million Veteran Program Core Acknowledgement for Publications**

3

**Last updated August 3, 2021**

**MVP Executive Committee**

- Co-Chair: J. Michael Gaziano, M.D., M.P.H. VA Boston Healthcare System, 150 S. Huntington Avenue, Boston, MA 02130
- Co-Chair: Sumitra Muralidhar, Ph.D. US Department of Veterans Affairs, 810 Vermont Avenue NW, Washington, DC 20420
- Rachel Ramoni, D.M.D., Sc.D., Chief VA Research and Development Officer US Department of Veterans Affairs, 810 Vermont Avenue NW, Washington, DC 20420
- Jean Beckham, Ph.D. Durham VA Medical Center, 508 Fulton Street, Durham, NC 27705
- Kyong-Mi Chang, M.D. Philadelphia VA Medical Center, 3900 Woodland Avenue, Philadelphia, PA 19104
- Philip S. Tsao, Ph.D. VA Palo Alto Health Care System, 3801 Miranda Avenue, Palo Alto, CA 94304
- James Breeling, M.D., Ex-Officio US Department of Veterans Affairs, 810 Vermont Avenue NW, Washington, DC 20420
- Grant Huang, Ph.D., Ex-Officio US Department of Veterans Affairs, 810 Vermont Avenue NW, Washington, DC 20420
- Juan P. Casas, M.D., Ph.D., Ex-Officio VA Boston Healthcare System, 150 S. Huntington Avenue, Boston, MA 02130

**MVP Program Office**

- Sumitra Muralidhar, Ph.D.

US Department of Veterans Affairs, 810 Vermont Avenue NW, Washington, DC 20420

- Jennifer Moser, Ph.D.

US Department of Veterans Affairs, 810 Vermont Avenue NW, Washington, DC 20420

**MVP Recruitment/Enrollment**

- MVP Cohort Management Director/Recruitment/Enrollment Director, Boston – Stacey B. Whitbourne, Ph.D.; Jessica V. Brewer, M.P.H.

VA Boston Healthcare System, 150 S. Huntington Avenue, Boston, MA 02130

- VA Central Biorepository, Boston – Mary T. Brophy M.D., M.P.H.; Donald E. Humphries, Ph.D.; Luis E. Selva, Ph.D.

VA Boston Healthcare System, 150 S. Huntington Avenue, Boston, MA 02130

- MVP Informatics, Boston – Nhan Do, M.D.; Shahpoor (Alex) Shayan, M.S.

VA Boston Healthcare System, 150 S. Huntington Avenue, Boston, MA 02130

- MVP Data Operations/Analytics, Boston – Kelly Cho, M.P.H., Ph.D.

VA Boston Healthcare System, 150 S. Huntington Avenue, Boston, MA 02130

- Director of Regulatory Affairs – Lori Churby, B.S.

VA Palo Alto Health Care System, 3801 Miranda Avenue, Palo Alto, CA 94304

- MVP Coordinating Centers
  - Cooperative Studies Program Clinical Research Pharmacy Coordinating Center, Albuquerque – Todd Connor, Pharm.D.; Dean P. Argyres, B.S., M.S.

New Mexico VA Health Care System, 1501 San Pedro Drive SE, Albuquerque, NM 87108

4

- Genomics Coordinating Center, Palo Alto – Philip S. Tsao, Ph.D.

VA Palo Alto Health Care System, 3801 Miranda Avenue, Palo Alto, CA 94304

- MVP Boston Coordinating Center, Boston - J. Michael Gaziano, M.D., M.P.H.

VA Boston Healthcare System, 150 S. Huntington Avenue, Boston, MA 02130

- MVP Information Center, Canandaigua – Brady Stephens, M.S.

Canandaigua VA Medical Center, 400 Fort Hill Avenue, Canandaigua, NY 14424

**MVP Science**

- Saiju Pyarajan Ph.D.

VA Boston Healthcare System, 150 S. Huntington Avenue, Boston, MA 02130

Philip S. Tsao, Ph.D.

VA Palo Alto Health Care System, 3801 Miranda Avenue, Palo Alto, CA 94304

- Data Core - Kelly Cho, M.P.H, Ph.D.

VA Boston Healthcare System, 150 S. Huntington Avenue, Boston, MA 02130

- VA Informatics and Computing Infrastructure (VINCI) – Scott L. DuVall, Ph.D.

VA Salt Lake City Health Care System, 500 Foothill Drive, Salt Lake City, UT 84148

- Data and Computational Sciences – Saiju Pyarajan, Ph.D.

VA Boston Healthcare System, 150 S. Huntington Avenue, Boston, MA 02130

- Statistical Genetics – Elizabeth Hauser, Ph.D.

Durham VA Medical Center, 508 Fulton Street, Durham, NC 27705

Yan Sun, Ph.D.

Atlanta VA Medical Center, 1670 Clairmont Road, Decatur, GA 30033

Hongyu Zhao, Ph.D.

West Haven VA Medical Center, 950 Campbell Avenue, West Haven, CT 06516

**Current MVP Local Site Investigators**

- Atlanta VA Medical Center (Peter Wilson, M.D.)

1670 Clairmont Road, Decatur, GA 30033

- Bay Pines VA Healthcare System (Rachel McArdle, Ph.D.)

10,000 Bay Pines Blvd Bay Pines, FL 33744

- Birmingham VA Medical Center (Louis Dellitalia, M.D.)

700 S. 19th Street, Birmingham AL 35233

- Central Western Massachusetts Healthcare System (Kristin Mattocks, Ph.D., M.P.H.)

421 North Main Street, Leeds, MA 01053

- Cincinnati VA Medical Center (John Harley, M.D., Ph.D.)

3200 Vine Street, Cincinnati, OH 45220 - Clement J. Zablocki

- VA Medical Center (Jeffrey Whittle, M.D., M.P.H.)

5000 West National Avenue, Milwaukee, WI 53295

- VA Northeast Ohio Healthcare System (Frank Jacono, M.D.)

10701 East Boulevard, Cleveland, OH 44106

- Durham VA Medical Center (Jean Beckham, Ph.D.)

508 Fulton Street, Durham, NC 27705

- Edith Nourse Rogers Memorial Veterans Hospital (John Wells., Ph.D.)

5

200 Springs Road, Bedford, MA 01730

- Edward Hines, Jr. VA Medical Center (Salvador Gutierrez, M.D.)

5000 South 5th Avenue, Hines, IL 60141

- Veterans Health Care System of the Ozarks (Kathrina Alexander, M.D.)

1100 North College Avenue, Fayetteville, AR 72703

- Fargo VA Health Care System (Kimberly Hammer, Ph.D.)

2101 N. Elm, Fargo, ND 58102

- VA Health Care Upstate New York (James Norton, Ph.D.)

113 Holland Avenue, Albany, NY 12208

- New Mexico VA Health Care System (Gerardo Villareal, M.D.)

1501 San Pedro Drive, S.E. Albuquerque, NM 87108

- VA Boston Healthcare System (Scott Kinlay, M.B.B.S., Ph.D.)

150 S. Huntington Avenue, Boston, MA 02130

- VA Western New York Healthcare System (Junzhe Xu, M.D.)

3495 Bailey Avenue, Buffalo, NY 14215-1199

- Ralph H. Johnson VA Medical Center (Mark Hamner, M.D.)

109 Bee Street, Mental Health Research, Charleston, SC 29401

- Columbia VA Health Care System (Roy Mathew, M.D.)

6439 Garners Ferry Road, Columbia, SC 29209

- VA North Texas Health Care System (Sujata Bhushan, M.D.)

4500 S. Lancaster Road, Dallas, TX 75216

- Hampton VA Medical Center (Pran Iruvanti, D.O., Ph.D.)

100 Emancipation Drive, Hampton, VA 23667

- Richmond VA Medical Center (Michael Godschalk, M.D.)

1201 Broad Rock Blvd., Richmond, VA 23249

- Iowa City VA Health Care System (Zuhair Ballas, M.D.)

601 Highway 6 West, Iowa City, IA 52246-2208

- Eastern Oklahoma VA Health Care System (River Smith, Ph.D.)

1011 Honor Heights Drive, Muskogee, OK 74401

- James A. Haley Veterans’ Hospital (Stephen Mastorides, M.D.)

13000 Bruce B. Downs Blvd, Tampa, FL 33612

- James H. Quillen VA Medical Center (Jonathan Moorman, M.D., Ph.D.)

Corner of Lamont & Veterans Way, Mountain Home, TN 37684

- John D. Dingell VA Medical Center (Saib Gappy, M.D.)

4646 John R Street, Detroit, MI 48201

- Louisville VA Medical Center (Jon Klein, M.D., Ph.D.)

800 Zorn Avenue, Louisville, KY 40206

- Manchester VA Medical Center (Nora Ratcliffe, M.D.)

718 Smyth Road, Manchester, NH 03104

- Miami VA Health Care System (Ana Palacio, M.D., M.P.H.)

1201 NW 16th Street, 11 GRC, Miami FL 33125

- Michael E. DeBakey VA Medical Center (Olaoluwa Okusaga, M.D.)

2002 Holcombe Blvd, Houston, TX 77030

- Minneapolis VA Health Care System (Maureen Murdoch, M.D., M.P.H.)

6

One Veterans Drive, Minneapolis, MN 55417

- N. FL/S. GA Veterans Health System (Peruvemba Sriram, M.D.)

1601 SW Archer Road, Gainesville, FL 32608

- Northport VA Medical Center (Shing Shing Yeh, Ph.D., M.D.)

79 Middleville Road, Northport, NY 11768

- Overton Brooks VA Medical Center (Neeraj Tandon, M.D.)

510 East Stoner Ave, Shreveport, LA 71101

- Philadelphia VA Medical Center (Darshana Jhala, M.D.)

3900 Woodland Avenue, Philadelphia, PA 19104

- Phoenix VA Health Care System (Samuel Aguayo, M.D.)

650 E. Indian School Road, Phoenix, AZ 85012

- Portland VA Medical Center (David Cohen, M.D.)

3710 SW U.S. Veterans Hospital Road, Portland, OR 97239

- Providence VA Medical Center (Satish Sharma, M.D.)

830 Chalkstone Avenue, Providence, RI 02908

- Richard Roudebush VA Medical Center (Suthat Liangpunsakul, M.D., M.P.H.)

1481 West 10th Street, Indianapolis, IN 46202

- Salem VA Medical Center (Kris Ann Oursler, M.D.)

1970 Roanoke Blvd, Salem, VA 24153

- San Francisco VA Health Care System (Mary Whooley, M.D.)

4150 Clement Street, San Francisco, CA 94121

- South Texas Veterans Health Care System (Sunil Ahuja, M.D.)

7400 Merton Minter Boulevard, San Antonio, TX 78229

- Southeast Louisiana Veterans Health Care System (Joseph Constans, Ph.D.)

2400 Canal Street, New Orleans, LA 70119

- Southern Arizona VA Health Care System (Paul Meyer, M.D., Ph.D.)

3601 S 6th Avenue, Tucson, AZ 85723

- Sioux Falls VA Health Care System (Jennifer Greco, M.D.)

2501 W 22nd Street, Sioux Falls, SD 57105

- St. Louis VA Health Care System (Michael Rauchman, M.D.)

915 North Grand Blvd, St. Louis, MO 63106

- Syracuse VA Medical Center (Richard Servatius, Ph.D.)

800 Irving Avenue, Syracuse, NY 13210

- VA Eastern Kansas Health Care System (Melinda Gaddy, Ph.D.)

4101 S 4th Street Trafficway, Leavenworth, KS 66048

- VA Greater Los Angeles Health Care System (Agnes Wallbom, M.D., M.S.)

11301 Wilshire Blvd, Los Angeles, CA 90073

- VA Long Beach Healthcare System (Timothy Morgan, M.D.)

5901 East 7th Street Long Beach, CA 90822

- VA Maine Healthcare System (Todd Stapley, D.O.)

1 VA Center, Augusta, ME 04330

- VA New York Harbor Healthcare System (Peter Liang, M.D., M.P.H.)

423 East 23rd Street, New York, NY 10010

- VA Pacific Islands Health Care System (Daryl Fujii, Ph.D.)

7

459 Patterson Rd, Honolulu, HI 96819

- VA Palo Alto Health Care System (Philip Tsao, Ph.D.)

3801 Miranda Avenue, Palo Alto, CA 94304-1290

- VA Pittsburgh Health Care System (Patrick Strollo, Jr., M.D.)

University Drive, Pittsburgh, PA 15240

- VA Puget Sound Health Care System (Edward Boyko, M.D.)

1660 S. Columbian Way, Seattle, WA 98108-1597

- VA Salt Lake City Health Care System (Jessica Walsh, M.D.)

500 Foothill Drive, Salt Lake City, UT 84148

- VA San Diego Healthcare System (Samir Gupta, M.D., M.S.C.S.)

3350 La Jolla Village Drive, San Diego, CA 92161

- VA Sierra Nevada Health Care System (Mostaqul Huq, Pharm.D., Ph.D.)

975 Kirman Avenue, Reno, NV 89502

- VA Southern Nevada Healthcare System (Joseph Fayad, M.D.)

6900 North Pecos Road, North Las Vegas, NV 89086

- VA Tennessee Valley Healthcare System (Adriana Hung, M.D., M.P.H.)

1310 24th Avenue, South Nashville, TN 37212

- Washington DC VA Medical Center (Jack Lichy, M.D., Ph.D.)

50 Irving St, Washington, D. C. 20422

- W.G. (Bill) Hefner VA Medical Center (Robin Hurley, M.D.)

1601 Brenner Ave, Salisbury, NC 28144

- White River Junction VA Medical Center (Brooks Robey, M.D.)

163 Veterans Drive, White River Junction, VT 05009

- William S. Middleton Memorial Veterans Hospital (Prakash Balasubramanian, M.D.)

2500 Overlook Terrace, Madison, WI 53705

#### **Materials and methods**

8

#### **Bioannotation and bioinformatic secondary analyses**

We conducted bioannotation and bioinformatic analyses to further characterize the loci identified by the GWAS meta-analysis of TUD-EUR). FUMA (version 1.3.5e)^1^ was used to explore the functional consequences of the genome-wide significant SNPs (**Supplementary Table 10**), which included ANNOVAR categories (the functional consequence of SNPs on genes), combined annotation dependent depletion scores, RegulomeDB scores, expression quantitative trait loci and chromatin states. The default external reference data for FUMA is described elsewhere.^1^

Gene-based analyses were performed with MAGMA (version 1.08). Genome-wide SNPs were first mapped to 19,532 protein-coding genes from Ensembl (build 85),^2^ and SNPs within each gene were jointly tested for association with TUD. We evaluated Bonferroni-corrected significance, adjusted for the number of genes (one-sided *p*<2.63E-06; **Supplementary Table 15**). Next, MAGMA gene-set analysis was performed using 18,868 curated gene sets and Gene Ontology terms obtained from the Molecular Signatures Database (version 7.0).^3^ We evaluated Bonferroni-corrected significance, adjusted for the number of gene sets (one-sided *p*<2.65E−06; **Supplementary Table 29**). A gene-property analysis tested the relationships between 54 tissue-specific gene expression profiles and gene associations while adjusting for the average expression of genes per tissue type as a covariate (**Supplementary Table 26**), and between brain gene expression profiles and gene associations across 11 brain tissues from BrainSpan^4,5^ (<http://www.brainspan.org/>). Gene expression values were log2-transformed average reads per kilobase million (**RPKM**) per tissue type (after replacing RPKM > 50 with 50) based on Genotype-Tissue Expression (**GTEx**) RNA-sequencing data (version 8.0). We evaluated Bonferroni-corrected significance, adjusted for the number of tested profiles (one-sided *p*<9.26E−04).

We used an extension of MAGMA: ‘Hi-C coupled MAGMA’ or ‘H-MAGMA’ (based on MAGMA version 1.08),^6^ to assign noncoding (intergenic and intronic) SNPs to cognate genes based on their chromatin interactions. Exonic and promoter SNPs were assigned to genes based on physical position. We used four Hi-C datasets provided with the software.^7^ We evaluated Bonferroni-corrected significance, adjusted for the number of tests within each of the four Hi-C datasets (one-sided *p*<9.44E-07; **Supplementary Table 16**).

9

We used S-MultiXcan v0.7.0 and S-PrediXcan v0.6.2 to test the association of TUD with gene expression in brain tissues. We used precomputed tissue weights from the GTEx database (version 8.0) as the reference dataset. As inputs, we used the TUD-EUR summary statistics, LD matrices of the SNPs (available at the PredictDB Data Repository; http://predictdb.org/) and transcriptome-tissue data related to 13 brain tissues: anterior cingulate cortex, amygdala, caudate basal ganglia, cerebellar hemisphere, cerebellum, cortex, frontal cortex, hippocampus, hypothalamus, nucleus accumbens basal ganglia, putamen basal ganglia, spinal cord and substantia nigra. We evaluated transcriptome-wide significance at the two-sided *p*< 2.31E−06, which was Bonferroni corrected for 21,610 unique tested genes (**Supplementary Table 17**).

**Phenome-wide association analyses**

***ABCD.*** We used the Rapid Imputation and COmputational PIpeLIne for Genome-Wide Association Studies (RICOPILI) to perform quality control (QC) on the 11,099 individuals with available ABCD Study phase 3.0 genotypic data, using RICOPILI’s default parameters.^8–12^ The 10,585 individuals who passed QC checks were matched to broad self-report racial groups using the ABCD Study parent survey. 6,787 parents/caregivers indicated that their child’s race was only “white”, and 5,561 of those individuals did not endorse any Hispanic ethnicity/origin. Further, we identified 1,675 parents/caregivers who indicated that their child’s race was only “black”, and 1,584 of those individuals did not endorse any Hispanic ethnicity/origin. After performing a second round of QC on these sub-samples, 5,556 non-Hispanic White and 1,584 non-Hispanic Black individuals were retained in the analyses. Principal component analysis (PCA) in RICOPILI was used to confirm the genetic ancestry of these individuals by mapping onto the 1000 Genomes reference panel, resulting in PCA-selected European- and African-ancestry subsets.

Each ancestry subset was then imputed to the TOPMed imputation reference panel.^38^ Imputation dosages were converted to best-guess hard-called genotypes, and only SNPs with R^2^ > 0.8 and MAF > 0.01 were kept for PGS analyses.

We used PRS-CS to calculate polygenic risk scores in the European ancestry subset of the ABCD sample, using effect sizes from the discovery GWAS summary statistics. We used the ‘auto’ function of PRS-CS, allowing the software to learn the global shrinkage parameter from the data. To maximize prediction in the African ancestry subset of ABCD, we used PRS-CSx’s ‘meta’ option to create polygenic risk scores that leveraged the larger sample size of the European ancestry version of the discovery GWAS by meta-analyzing those weights along with weights from the smaller, ancestry-matched discovery GWAS.

After deriving SNP weights using PRS-CS and PRS-CS, we then used PLINK 1.9’s --score command to produce PGS in the ABCD sample. All subsequent analyses were performed using R Statistical Software.^12^ We scaled the PGS to a mean of zero and standard deviation of one before including them in regression models.

**Supplementary Figures**

11

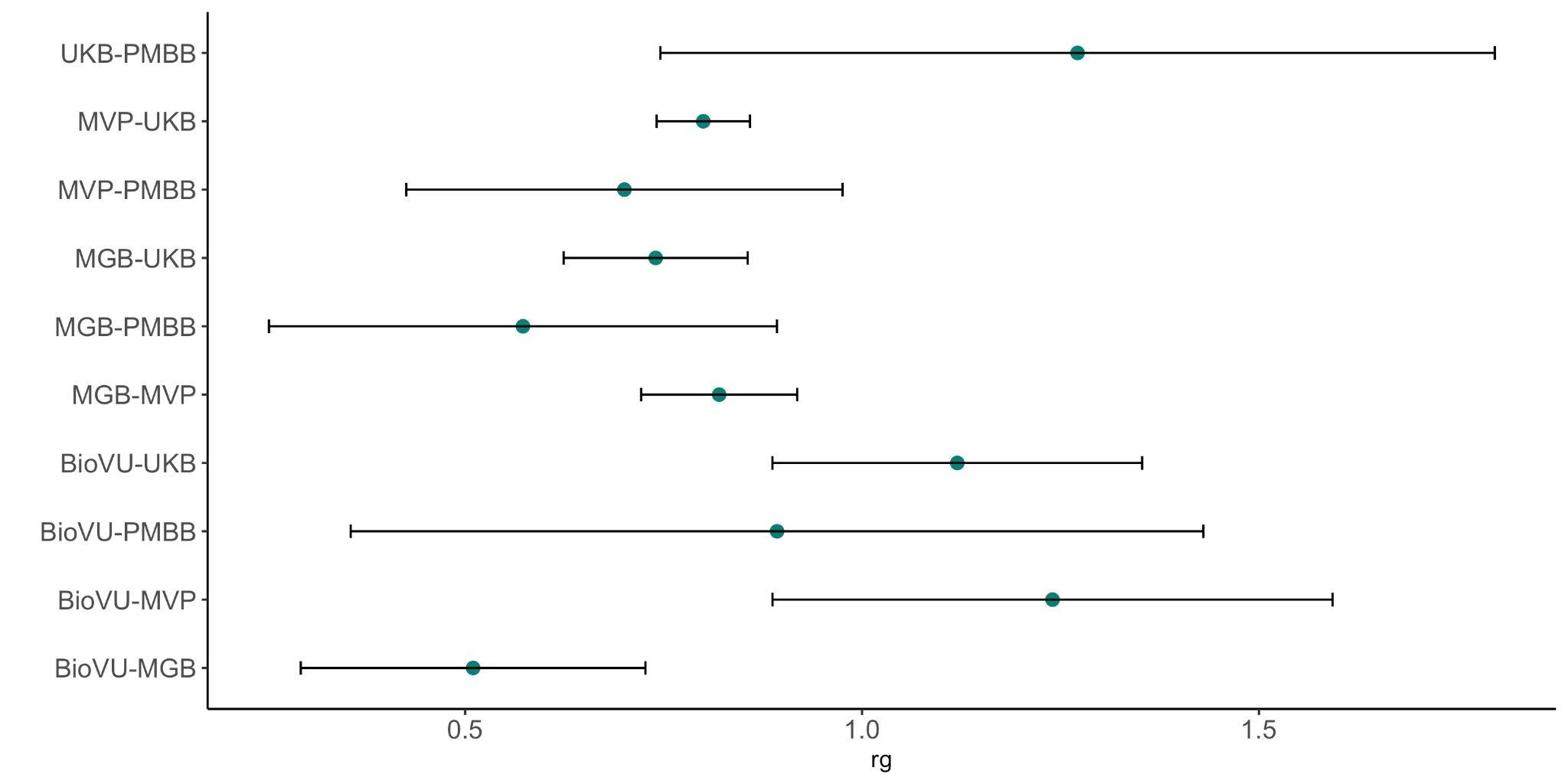

**Supplementary Figure 1.** LDSC genetic correlations for TUD between all EUR sites with confidence intervals.

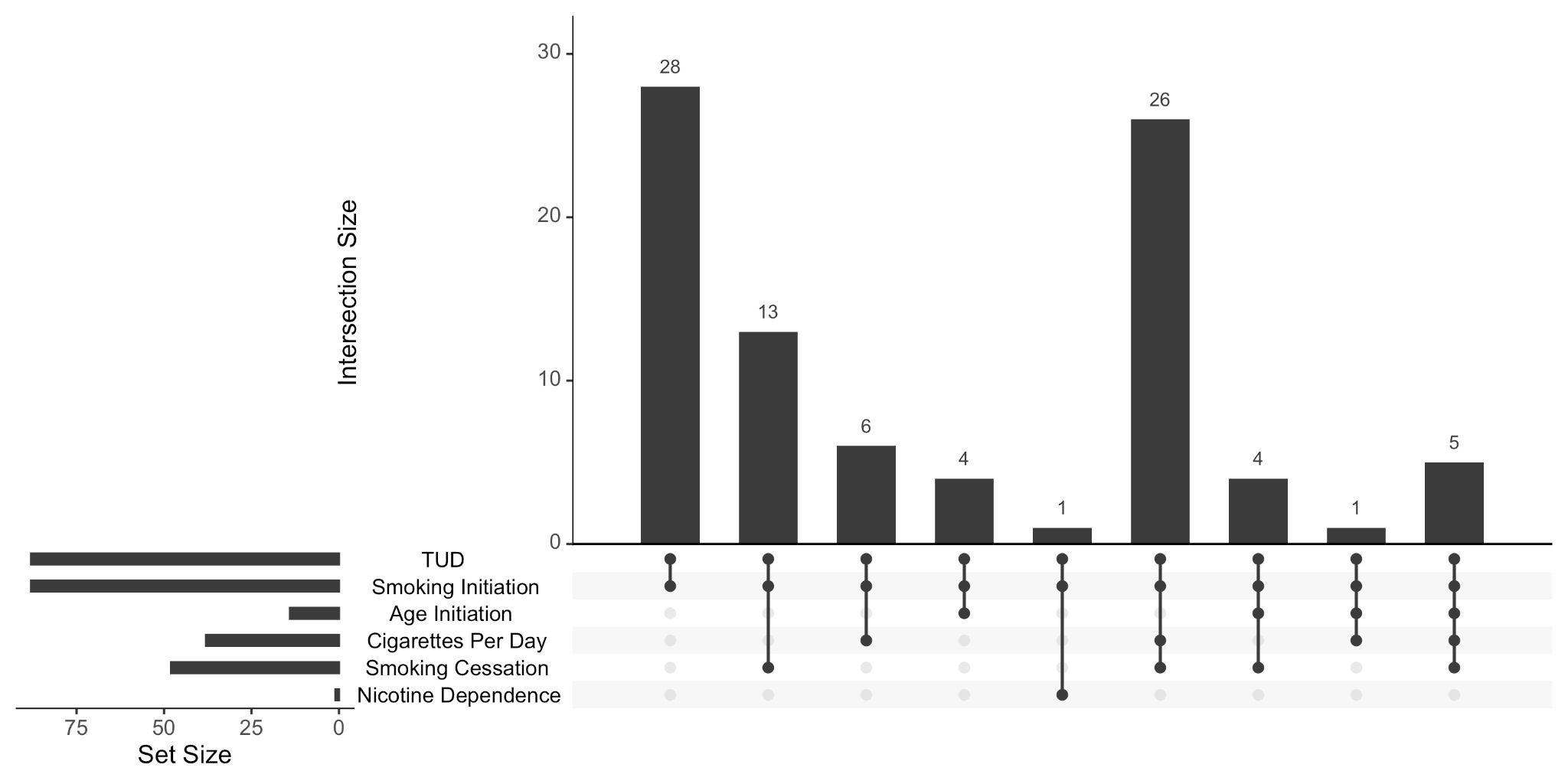

12

**Supplementary Figure 2.** Multi-ancestry TUD meta-analysis replicates previously known associations with smoking traits, from initiation, to consumption, to cessation (GSCAN2), to dependence (FTND). Shown are only the loci which overlap between the current study and previous smoking GWAS.

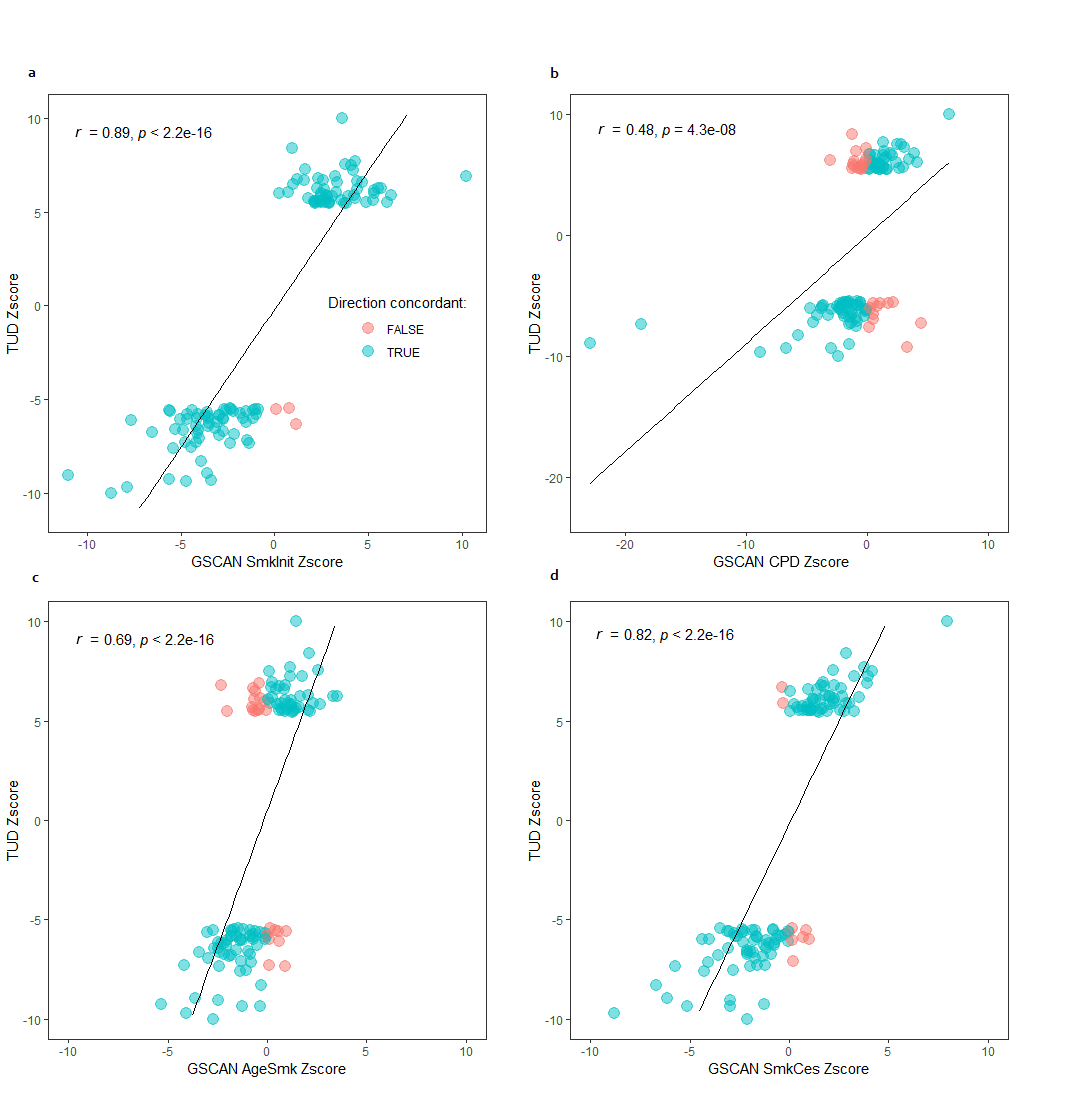

13

**Supplementary Figure 3.** Comparison of effect sizes across TUD-multi and smoking GSCAN2 GWASs. The magnitude and direction of the effect sizes of TUD lead SNPs and GSCAN smoking behaviors (smoking initiation, SmkInit; age of smoking initiation, AgeSmk; Cigarettes per day, CPD; smoking cessation, SmkCess). The results show significant (*p* < 2.2E-05) high to moderate correlations (*r*) between effect sizes (Z scores) of TUD lead SNPs and GSCAN2 SNPs for SmkInit (*r* = 0.89, **a**), CPD (*r* = 0.48, **b**), AgeSmk (reverse coded, *r* = 0.69, **c**) and SmkCes (*r* = 0.82, **d**).

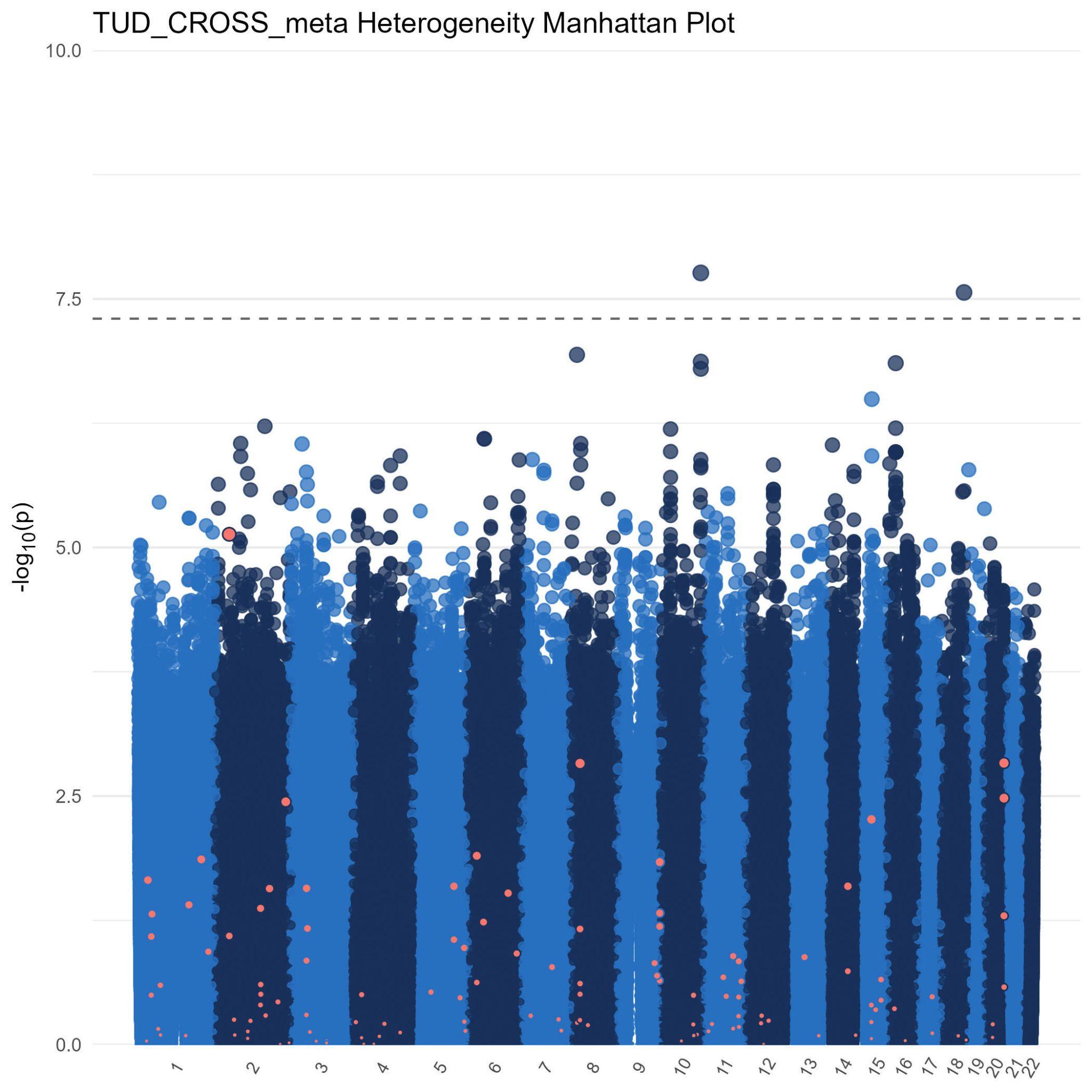

14
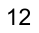

**Supplementary Figure 4.** Heterogeneity plot of TUD-multi meta-analysis indicates no signs of heterogeneity (*I^2^*).

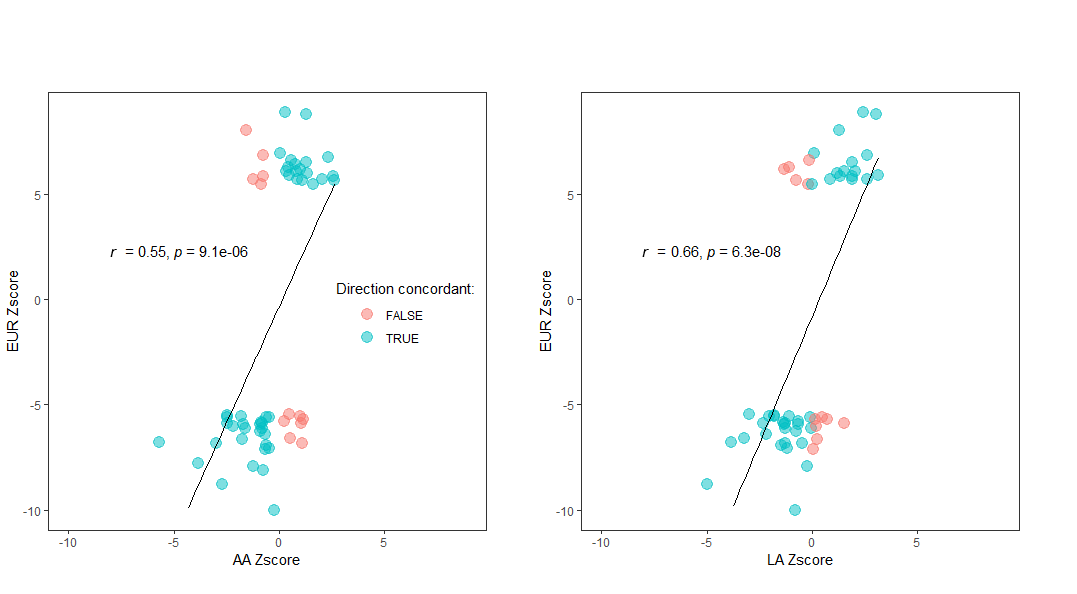
**Supplementary Figure 5.** Effect-effect plot showing the effect size of the TUD lead SNPs across ancestries (AA, African American, left; LA, Latin American, right).

15

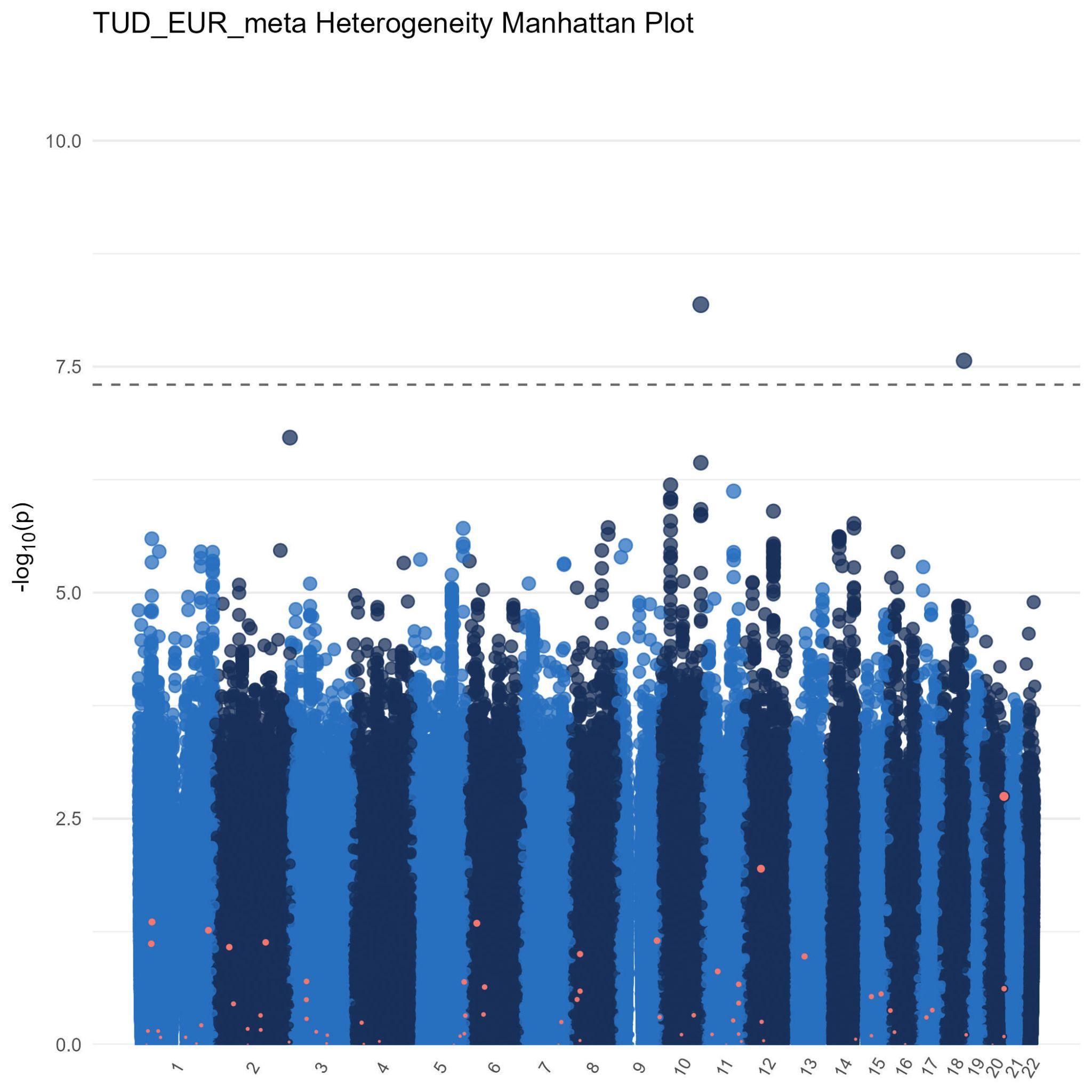

16

**Supplementary Figure 6.** Heterogeneity plot of TUD-EUR meta-analysis indicates no signs of heterogeneity (*I^2^*).

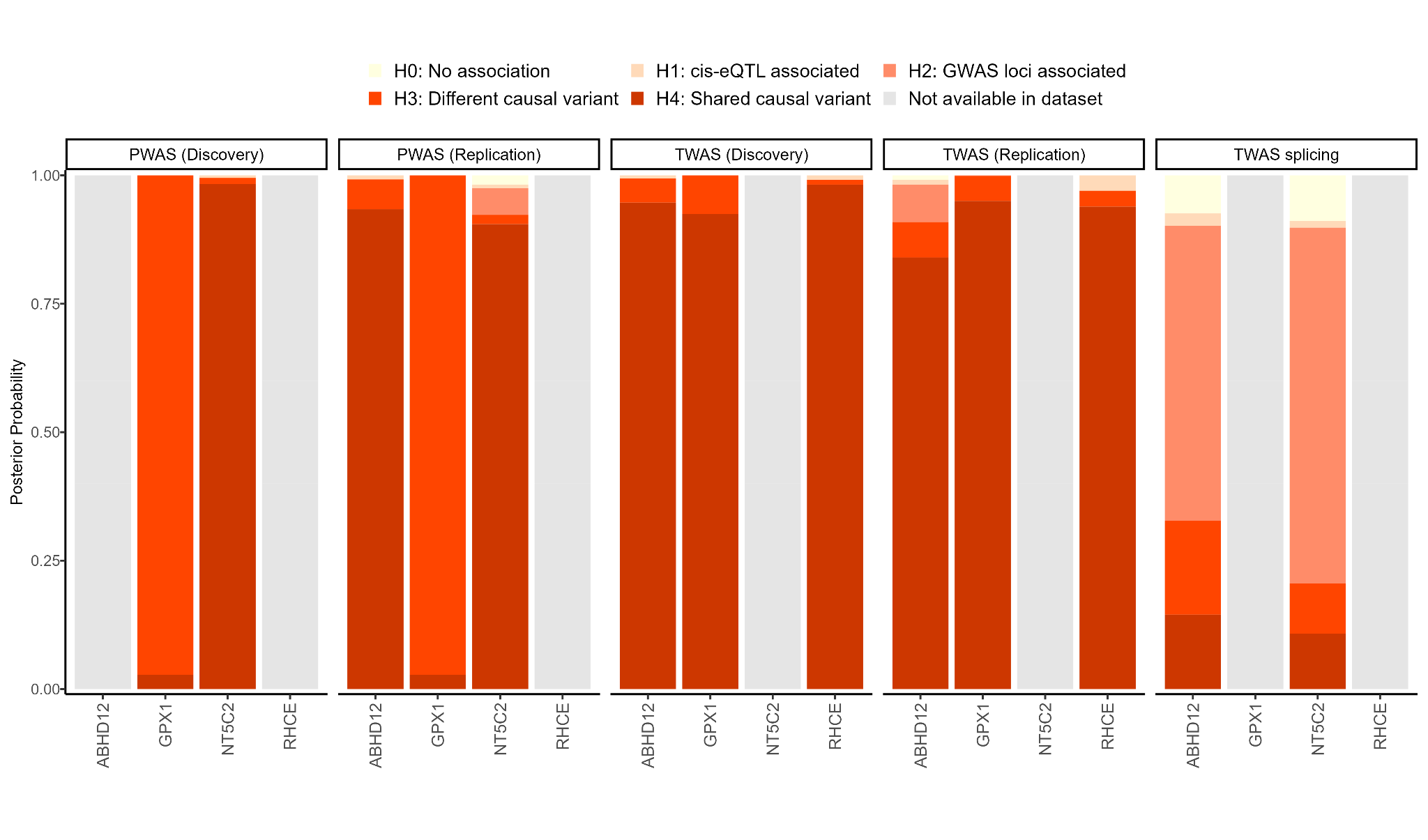

17

**Supplementary Figure 7.** Genes identified via TWAS and PWAS analyses revealed levels of regulation of TUD risk genes either via gene regulation or protein abundance, respectively, in the brain.

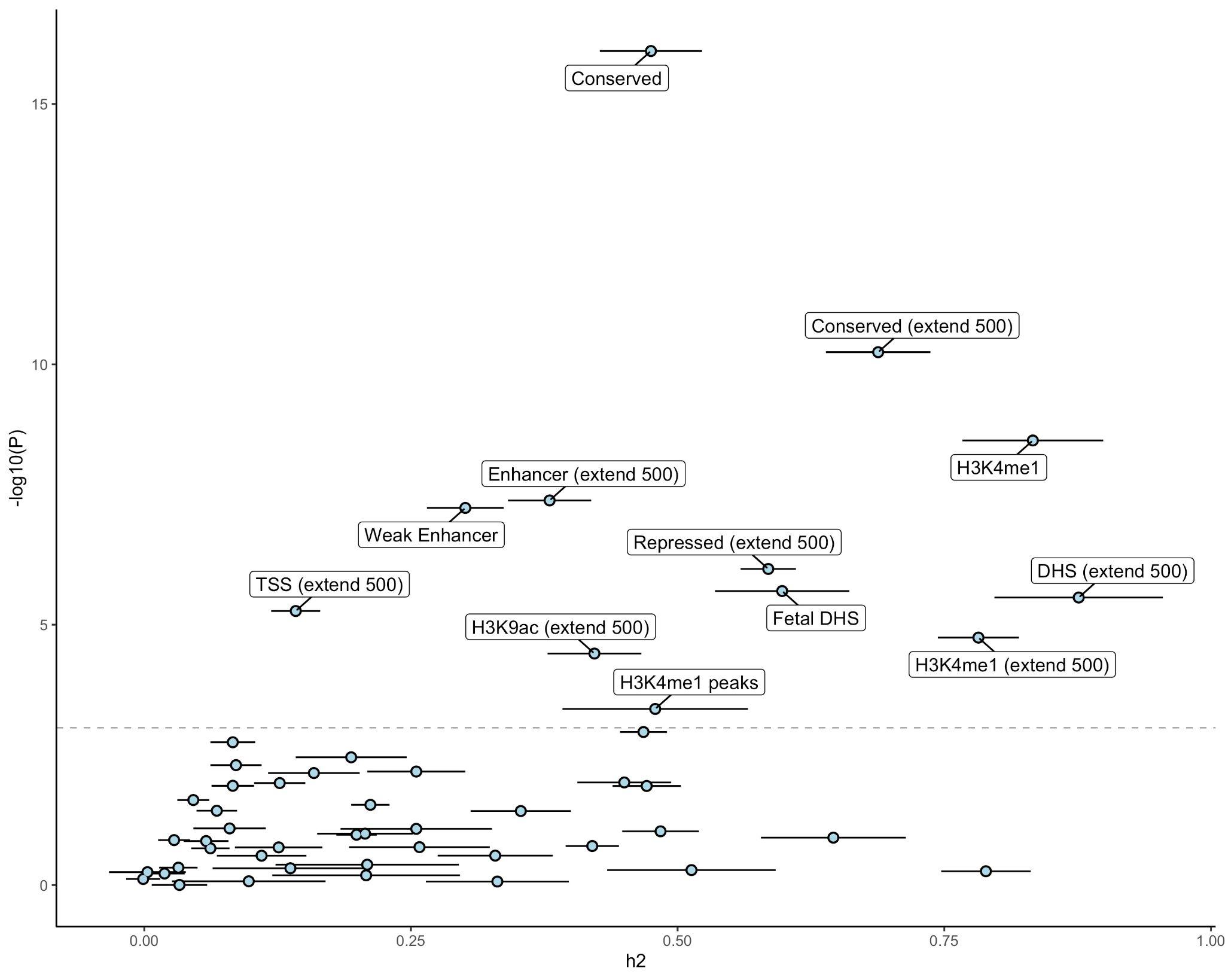

18

**Supplementary Figure 8.** Partitioned heritability in LDSC to evaluate enrichment of the GWS findings in over 50 functional genomic annotations (and across tissues). In the baseline LDSC model, conserved and regulatory functional annotations were significantly enriched.

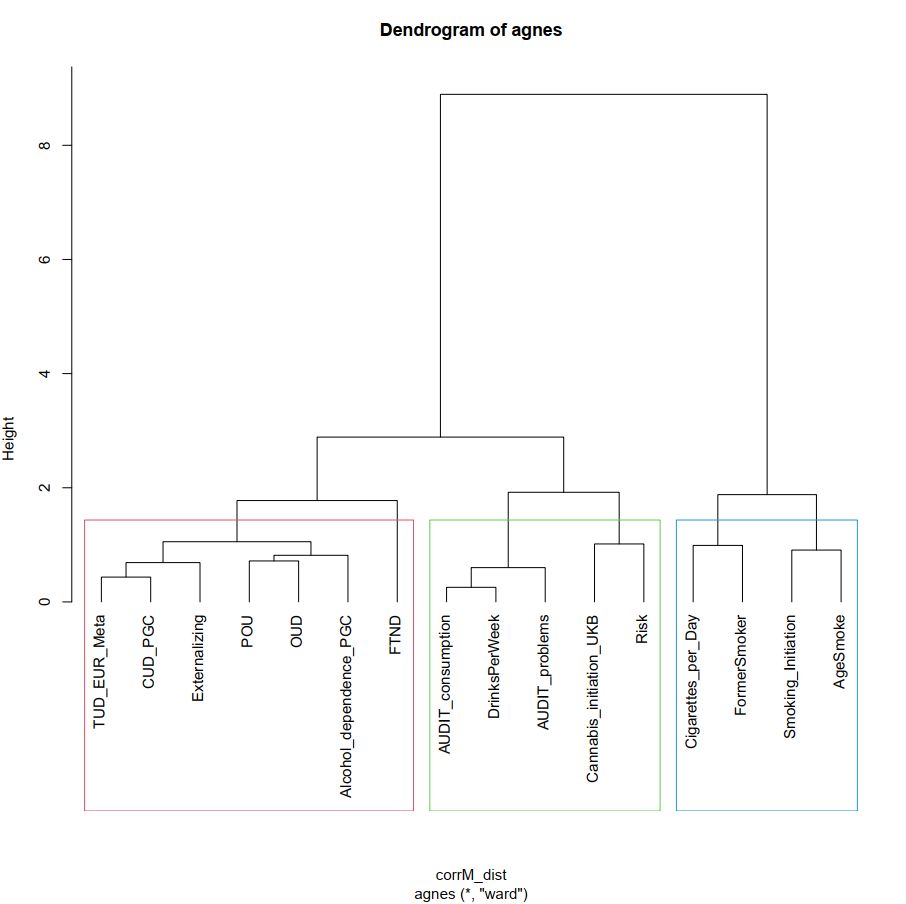

19

**Supplementary Figure 9.** Hierarchical Clustering Analysis with Agglomerative Nesting (HCA with AGNES) dendrogram**.** Input was the pairwise genetic correlation matrix. Boxes represent the division in branches needed to divide substance use and use disorder traits. Height (y-axis) represents distance metrics between the clusters. *Abbreviations.* TUD_EUR_Meta, current study; FTND, Fagerström Test for Nicotine Dependence, POU, Problematic Opioid Use; CUD_PGNC, Cannabis Use Disorder; OUD, Opioid Use Disorder (MVP); Risk, General Risk Tolerance; AgeSmoke, Age of Initiation.

**REFERENCES**

20

1.	Watanabe, K., Taskesen, E., van Bochoven, A. & Posthuma, D. Functional mapping and annotation of genetic associations with FUMA. *Nat. Commun.* **8**, 1826 (2017).

2.	Cunningham, F. *et al.* Ensembl 2022. *Nucleic Acids Res.* **50**, D988–D995 (2021).

3.	Leeuw, C. A. de, Mooij, J. M., Heskes, T. & Posthuma, D. MAGMA: Generalized Gene-Set Analysis of GWAS Data. *PLOS Comput. Biol.* **11**, e1004219 (2015).

4.	Gulsuner, S. *et al.* Spatial and temporal mapping of de novo mutations in schizophrenia to a fetal prefrontal cortical network. *Cell* **154**, 518–529 (2013).

5.	Kang, H. J. *et al.* Spatio-temporal transcriptome of the human brain. *Nature* **478**, 483–489 (2011).

6.	Sey, N. Y. A. *et al.* A computational tool (H-MAGMA) for improved prediction of brain-disorder risk genes by incorporating brain chromatin interaction profiles. *Nat. Neurosci.* **23**, 583–593 (2020).

7.	Schmitt, A. D. *et al.* A Compendium of Chromatin Contact Maps Reveals Spatially Active Regions in the Human Genome. *Cell Rep.* **17**, 2042–2059 (2016).

8.	Lam, M. *et al.* RICOPILI: Rapid Imputation for COnsortias PIpeLIne. *Bioinformatics* **36**, 930–933 (2020).

9.	Taliun, D. *et al.* Sequencing of 53,831 diverse genomes from the NHLBI TOPMed Program. *Nature* **590**, 290–299 (2021).

10. Ge, T., Chen, C.-Y., Ni, Y., Feng, Y.-C. A. & Smoller, J. W. Polygenic prediction via Bayesian regression and continuous shrinkage priors. *Nat. Commun.* **10**, 1776 (2019).

11. Ruan, Y. *et al.* Improving Polygenic Prediction in Ancestrally Diverse Populations. *medRxiv* 21 (2021) doi:https://doi.org/10.1101/2020.12.27.20248738.

12. Chang, C. C. *et al.* Second-generation PLINK: rising to the challenge of larger and richer datasets. *GigaScience* **4**, 7 (2015).
